# Large-scale Multi-omic Analysis of COVID-19 Severity

**DOI:** 10.1101/2020.07.17.20156513

**Authors:** Katherine A. Overmyer, Evgenia Shishkova, Ian J. Miller, Joseph Balnis, Matthew N. Bernstein, Trenton M. Peters-Clarke, Jesse G. Meyer, Qiuwen Quan, Laura K. Muehlbauer, Edna A. Trujillo, Yuchen He, Amit Chopra, Hau C. Chieng, Anupama Tiwari, Marc A. Judson, Brett Paulson, Dain R. Brademan, Yunyun Zhu, Lia R. Serrano, Vanessa Linke, Lisa A. Drake, Alejandro P. Adam, Bradford S. Schwartz, Harold A. Singer, Scott Swanson, Deane F. Mosher, Ron Stewart, Joshua J. Coon, Ariel Jaitovich

**Affiliations:** National Center for Quantitative Biology of Complex Systems, Madison, WI 53562, USA; Morgridge Institute for Research, Madison, WI 53562, USA; Department of Biomolecular Chemistry, University of Wisconsin, Madison, WI 53562, USA; Division of Pulmonary and Critical Care Medicine, Albany Medical Center, Albany, NY, USA; Department of Molecular and Cellular Physiology, Albany Medical College, Albany, NY, USA; Department of Chemistry, University of Wisconsin, Madison, WI 53562, USA; Division of Sleep Medicine, Albany Medical Center, Albany, NY, USA; Department of Ophthalmology, Albany Medical College, Albany, NY, USA

**Author notes:** to whom correspondence should be addressed A.J., J.J.C. these authors contributed equally (Katherine A. Overmyer, Evgenia Shishkova, Ian J. Miller, Joseph Balnis). these authors contributed equally as co-second authors (Matthew N. Bernstein, Trenton M. Peters-Clarke, Jesse G. Meyer).

## Abstract

We performed RNA-Seq and high-resolution mass spectrometry on 128 blood samples from COVID-19 positive and negative patients with diverse disease severities. Over 17,000 transcripts, proteins, metabolites, and lipids were quantified and associated with clinical outcomes in a curated relational database, uniquely enabling systems analysis and cross-ome correlations to molecules and patient prognoses. We mapped 219 molecular features with high significance to COVID-19 status and severity, many involved in complement activation, dysregulated lipid transport, and neutrophil activation. We identified sets of covarying molecules, e.g., protein gelsolin and metabolite citrate or plasmalogens and apolipoproteins, offering pathophysiological insights and therapeutic suggestions. The observed dysregulation of platelet function, blood coagulation, acute phase response, and endotheliopathy further illuminated the unique COVID-19 phenotype. We present a web-based tool (covid-omics.app) enabling interactive exploration of our compendium and illustrate its utility through a comparative analysis with published data and a machine learning approach for prediction of COVID-19 severity.

## INTRODUCTION

As of July 2020, the COVID-19 pandemic has caused over 500,000 deaths worldwide, primarily due to complications from SARS-CoV-2-associated acute respiratory distress syndrome (ARDS) <u>(Guan et al., 2020)</u>. The clinical course of COVID-19 infection is highly variable, ranging from an asymptomatic state to a life-threatening infection. Recent evidence indicates that disease severity predominantly depends on host, rather than viral factors <u>(Zhang et al., 2020a)</u>, supporting the need to better understand the individuals’ response at a molecular level. While rapidly accumulating evidence indicates that distinct genetic <u>(Ellinghaus et al., 2020)</u>, physiological <u>(Gattinoni et al., 2020a)</u>, pathological <u>(Fox et al., 2020a)</u>, and clinical <u>(Richardson et al., 2020a)</u> signatures differentiate patients with and without COVID-19-driven ARDS, more clarity on the molecular basis explaining the observed differences is needed.

It is widely accepted that clinical syndromes of non-COVID-19 ARDS and sepsis result from an aggregation of different patient subgroups with distinct molecular signatures and responses to standardized treatments <u>(Reddy et al., 2020)</u>. For example, in non-COVID-19 ARDS patients, a relatively hyper-inflammatory phenotype is associated with higher mortality than a hypo-inflammatory state <u>(Calfee et al., 2014)</u>. Information on the association of the inflammatory landscape with COVID-19 patients’ outcomes is less clear <u>(Sinha et al., 2020)</u>. Moreover, even though leukocytes from patients with severe infections demonstrate an association between a robust inflammatory transcriptome and better prognosis <u>(Davenport et al., 2016a)</u>, it remains unclear to what extent the leukocyte gene expression profiles differ between patients with and without COVID-19. For these reasons, gaining further insight into the complex molecular environment of COVID-19 patients could facilitate identification of potential therapeutic targets of immunomodulation leading to better outcomes. Beyond the typical inflammatory response, COVID-19 patients demonstrate profound coagulation dysregulation <u>(Zhang et al., 2020c)</u>. A sensitive marker of fibrinogen degradation, D-dimer, is found consistently elevated in severe cases of COVID-19 <u>(Guan et al., 2020; Richardson et al., 2020b)</u>. In addition, data from autopsies demonstrate microvascular clotting <u>(Fox et al., 2020b)</u>, and clinical observations suggest that anticoagulation therapies may lower mortality of COVID-19-associated ARDS <u>(Paranjpe et al., 2020)</u>.

Despite the rapid global scientific response to this new disease, few studies have investigated the broad molecular level reorganization that drives the host COVID-19 viral response. Technologies for deep sequence analysis of nucleic acids (i.e., transcriptomics) are broadly available. High-resolution mass spectrometry can provide similar quantitative data for large-scale protein, metabolite, and lipid measurements. With the supposition that broad profiling across these various planes of biomolecular regulation could allow for a holistic view of disease pathophysiology, we sought to leverage these technologies on a large number of patients.

Accordingly, we conducted a cohort study involving 128 patients with and without COVID-19 diagnosis. To ensure that we generated molecular profiles that could illuminate the COVID-19 pathological signature, protein, metabolite, and lipid profiles were measured from blood plasma. Additionally, leukocytes derived from patient blood samples were isolated for RNA sequencing. Using state-of-the-art sequencing and mass spectrometric technologies we identified and quantified over 17,000 transcripts, proteins, metabolites, and lipids across these 128 patient samples. The abundances of these biomolecules were then correlated with a range of clinical data and patient outcomes to create a rich molecular resource for COVID-19 host response to be made available to the biomedical research community.

Here we leverage this resource to examine the pathophysiology of COVID-19, identify potential therapeutic opportunities, and facilitate accurate predictions of patient outcome. Briefly, we found 219 biomolecules which were highly correlated with COVID-19 status and severity. Tapping into the ability of our multi-omic method to uncover functional connections between different biomolecule classes, we discover sets of covarying molecules that shed light on disease mechanisms and provide therapeutic opportunities. As partially uncovered by earlier reports, our findings reveal numerous dysregulated biological processes in COVID-19, including complement system activation, lipid transport, vessel damage, platelet activation and degranulation, blood coagulation, and acute phase response. Given the focus of our study on plasma, our data provide unique insight into the COVID-19 hypercoagulation phenotype. This rich molecular compendium is made available through a freely accessible web resource (covid-omics.app) to aid the current global efforts to study this disease. We also present two application examples that leverage this resource: (1) a comparative analysis of these transcriptomics data and published data on classical ARDS, where we reveal distinct molecular signatures of COVID-19-instigated ARDS and non-COVID-19 ARDS; and (2) development of a disease severity predictive model based on all omics data. While patients’ comorbidities are powerfully associated with COVID-19 outcomes, our mulit-omics-based predictive model significantly improved COVID-19-severity predictions over the standardized clinical Charlson comorbidity score <u>(Richardson et al., 2020b)</u>.

## RESULTS

### Sample cohort and experimental design

From 6 April 2020 through 1 May 2020, we collected blood samples from 128 adult patients admitted to Albany Medical Center in Albany, NY for moderate to severe respiratory issues presumably related to infection with SARS-CoV-2. In addition to collection of various clinical data (**Table 1**), a blood sample was obtained at the time of enrollment. Patients who tested positive (n = 102) and negative (n = 26) for the virus were assigned to the COVID-19 and non-COVID-19 groups, respectively (see **Methods** for enrollment details). Females comprised 37.3% and 50.0% of the COVID-19 and non-COVID-19 groups, respectively. The average age of patients was similar – 61.3 and 63.1 years in the COVID-19 group (females and males, respectively; p-value = 0.56) – with some statistically insignificant differences between females and males in the non-COVID-19 group (59.5 and 67.0 years, respectively; p-value = 0.25) (**Figure 1a**). The average number of days hospitalized before study enrollment was 3 and 1 for the COVID-19 and non-COVID-19 groups, respectively (**Table 1**). The COVID-19 group was more racially diverse than the non-COVID-19 group, with white individuals comprising only 46% of the total (vs. 80% of the non-COVID-19 control group); these demographics are consistent with the reported racial and ethnic health disparities of COVID-19 <u>(Webb Hooper et al., 2020)</u>.

**Table 1:** Demographics and baseline characteristics of COVID-19 and non-Covid 19 patients in ICU and non-ICU setting.

| Variables | Total<br>n=102 | COVID-19 |  | Total<br>n=26 | non-COVID-19 |  |
| --- | --- | --- | --- | --- | --- | --- |
|  |  | non-ICU<br>n=51 | ICU<br>n=51 |  | non-ICU<br>n=10 | ICU<br>n=16 |
| Days Admitted Pre-Enrollment (IQR) | 3.37 (1-5) | 2.78 (1-3) | 3.96 (1-6) | 0.97 (1-1) | 0.9 (0.8-1) | 0.94 (1-1) |
| Sex - n (%) |  |  |  |  |  |  |
| Male | 64 (62.7%) | 30 (58.8%) | 34 (66.7%) | 13 (50%) | 4 (40%) | 9 (56%) |
| Female | 38 (37.3%) | 21 (41.2%) | 17 (33.3%) | 13 (50%) | 6 (60%) | 7 (44%) |
| Age-year |  |  |  |  |  |  |
| Mean (IQR) | 61.3 (50.0-74.3) | 59.7 (49.0-80.0) | 62.9 (55.0-73.0) | 63.8 (52.3-76.8) | 60.4 (47.3-74.0) | 66 (55.3-80.3) |
| Ethnicity - n (%) |  |  |  |  |  |  |
| White | 46 (45.1%) | 28 (54.9%) | 18 (35.3%) | 21 (80.8%) | 8 (80%) | 13 (81.2%) |
| Black | 11 (10.8%) | 5 (9.8%) | 6 (11.8%) | 4 (15.4%) | 2 (20%) | 2 (12.5%) |
| Asian | 2 (1.9%) | 0 (0%) | 2 (3.9%) | 0 (0%) | 0 (0%) | 0 (0%) |
| Hispanic | 21 (20.6%) | 7 (13.7%) | 14 (27.5%) | 1 (3.8%) | 0 (0%) | 1 (6.3%) |
| Other | 22 (21.6%) | 11 (21.6%) | 11 (21.6%) | 0 (0%) | 0 (0%) | 0 (0%) |
| BMI, kg/m2 Mean (IQR) □ | 30.39 (25.30-32.24) | 29.84 (26.09-32.37) | 30.92 (24.50-32.05) | 30.36 (26.53-33.10) | 27.20 (23.68-30.38) | 32.34 (26.98-37.67) |
| Severity Indexes (IQR) |  |  |  |  |  |  |
| Charlson comorbidity index | 3.3 (1-5) | 3.16 (1-5) | 3.49 (2-5) | 4.35 (2-6) | 3.3 (1-5) | 5 (3-7) |
| APACHEII | N/A | N/A | 21.6 (15-27) | N/A | N/A | 20.6 (12-26) |
| SOFA | N/A | N/A | 8.2 (6-11) | N/A | N/A | 8.6 (3-11) |
| SAPSII | N/A | N/A | 51.8 (45-62) | N/A | N/A | 47.6 (33-65) |
| Biomarkers (IQR) |  |  |  |  |  |  |
| Ferritin (ng/mL) | 938.9 (301.8-1203.8) | 782.6 (206.0-934.5) | 1076.9 (378.0-1294.0) | 250.5 (80.5-382.5) | 205.3 (58.0-411.0) | 285.7 (92.0-438.5) |
| C-Reactive protein (mg/L) | 140.9 (52.0-204.3) | 120.6 (44.7-155.0) | 158.9 (61.7-248.3) | 73.8 (20.0-110.2) | 34.7 (8.9-56.8) | 99.8 (37.8-175.2) |
| D-dimer (mg/L FEU) | 11.7 (1.0-12.8) | 2.3 (0.6-1.73) | 18.6 (1.7-21.6) | 5.3 (0.5-4.6) | 5.2 (0.4-1.9) | 5.5 (0.6-10.2) |
| Procalcitonin (ng/mL) | 3.2 (0.2-1.8) | 1.7 (0.2-1.0) | 4.4 (0.3-2.3) | 2.1 (0.2-0.7) | 2.2 (0.1-3.4) | 2.1 (0.3-1.21) |
| Lactate (mmol/L) | 1.2 (0.9-1.5) | 1.2 (0.9-1.4) | 1.3 (0.9-1.5) | 2.1 (0.9-2.5) | 1.2 (0.8-1.5) | 2.53 (0.9-3.4) |
| Fibrinogen (mg/dL) | 543.6 (413.0-667.0) | 559.3 (420.0-703.0) | 531.7 (391.5-663.0) | 362.3 (257.3-550.0) | 348.0 (256.75-441.5) | 373 (257.3-572.0) |
| Albumin (mg/L) | 2.9 (2.6-3.3) | 3.2 (2.9-3.5) | 2.7 (2.4-2.9) | 3.4 (2.9-3.8) | 3.8 (3.4-4.1) | 3.19 (2.6-3.8) |
| Hemogram (IQR) |  |  |  |  |  |  |
| White blood cells (K/uL) | 10.8 (6.1-12.5) | 7.1 (4.9-8.5) | 14.4 (8.4-15.4) | 12.7 (7.2-17.3) | 8.3 (6.7-9.7) | 15.4 (8.2-20.9) |
| Hemoglobin (g/dL) | 11.2 (9.7-12.6) | 11.6 (10.2-13.0) | 10.7 (9.4-12.1) | 12.4 (9.9-14.7) | 12.8 (10.45-14.85) | 12.3 (9.6-14.5) |
| Mean corpuscular volume (fL) | 87.1 (84.5-93.7) | 88.0 (85.6-94.2) | 86.2 (82.5-93.0) | 92.3 (88.6-95.4) | 91.2 (87.2-94.6) | 93.0 (89.4-97.8) |
| Platelet (K/uL) | 266.0 (192.5-320.5) | 269.2 (209.0-334) | 262.8 (187.0-317.0) | 203.5 (151.8-247.8) | 228.1 (163.5-278.0) | 188.2 (127.5-229.5) |
| Neutrophils (%) | 76.2 (68.5-86.0) | 69.7 (61.0-82.0) | 82.8 (80.0-90.0) | 77.7 (74.0-87.0) | 73.1 (58.8-82.5) | 80.5 (79.25-89.25) |
| Lymphocytes (%) | 13.8 (5.0-18.5) | 19.4 (9.0-26.0) | 8.3 (4.0-11.0) | 12.7 (6.0-18.0) | 16.9 (7.0-26.0) | 10.1 (4.3-10.8) |
| Monocytes (%) | 7.1 (4.0-9.0) | 8.8 (6.0-11.0) | 5.5 (3.0-8.0) | 8.0 (4.0-9.3) | 7.7 (4.0-10.3) | 8.2 (4.0-9.0) |
| Eosinophils (%) | 0.8 (0.0-1.0) | 1.1 (0.0-1.0) | 0.5 (0.0-1.0) | 1.0 (0.0-1.25) | 1.8 (0.0-3.3) | 0.44 (0.0-1.0) |
| Respiratory parameters |  |  |  |  |  |  |
| PaO2/FiO2 Ratio | N/A | N/A | 161.6 (98-211) | N/A | N/A | 149.4 (73-184) |
| Positive-end expiratory pressure (cmH2O) | N/A | N/A | 10.8 (10-12) | N/A | N/A | 6.6 (73-184) |
| Inspiratory Plateau (cmH2O) | N/A | N/A | 22.8 (19.7-25.3) | N/A | N/A | 23.9 (19.8-28.8) |
| Treatment - n (%) |  |  |  |  |  |  |
| Renal Replacement Therapy (Pre-enrollment) | 12 (11.8%) | 3 (5.9%) | 9 (17.6%) | 3 (11.5%) | 0 (0%) | 3 (18.8%) |
| Hydroxychloroquine | 87 (85.3%) | 43 (84.3%) | 44 (86.3%) | 0 (0%) | 0 (0%) | 0 (0%) |
| Antibiotics | 98 (96.1%) | 47 (92.2%) | 51 (100%) | 16 (61.5%) | 3 (30.0%) | 13 (81.3%) |
| Antiviral | 1 (0.98%) | 0 (0%) | 1 (1.9%) | 0 (0%) | 0 (0%) | 0 (0%) |
| IL6- Antagonist | 4 (3.9%) | 1 (1.9%) | 2 (3.9%) | 0 (0%) | 0 (0%) | 0 (0%) |
| Convalescent Plasma | 26 (25.5%) | 8 (15.7%) | 18 (35.3%) | 0 (0%) | 0 (0%) | 0 (0%) |
| Steroid | 46 (45.1%) | 12 (23.5%) | 34 (66.7%) | 4 (15.4%) | 1 (10.0%) | 3 (18.8%) |
| Therapeutic Anticoagulation | 37 (36.3%) | 2 (3.9%) | 35 (68.6%) | 8 (30.8%) | 1 (10.0%) | 7 (43.8%) |

**Figure 1.**
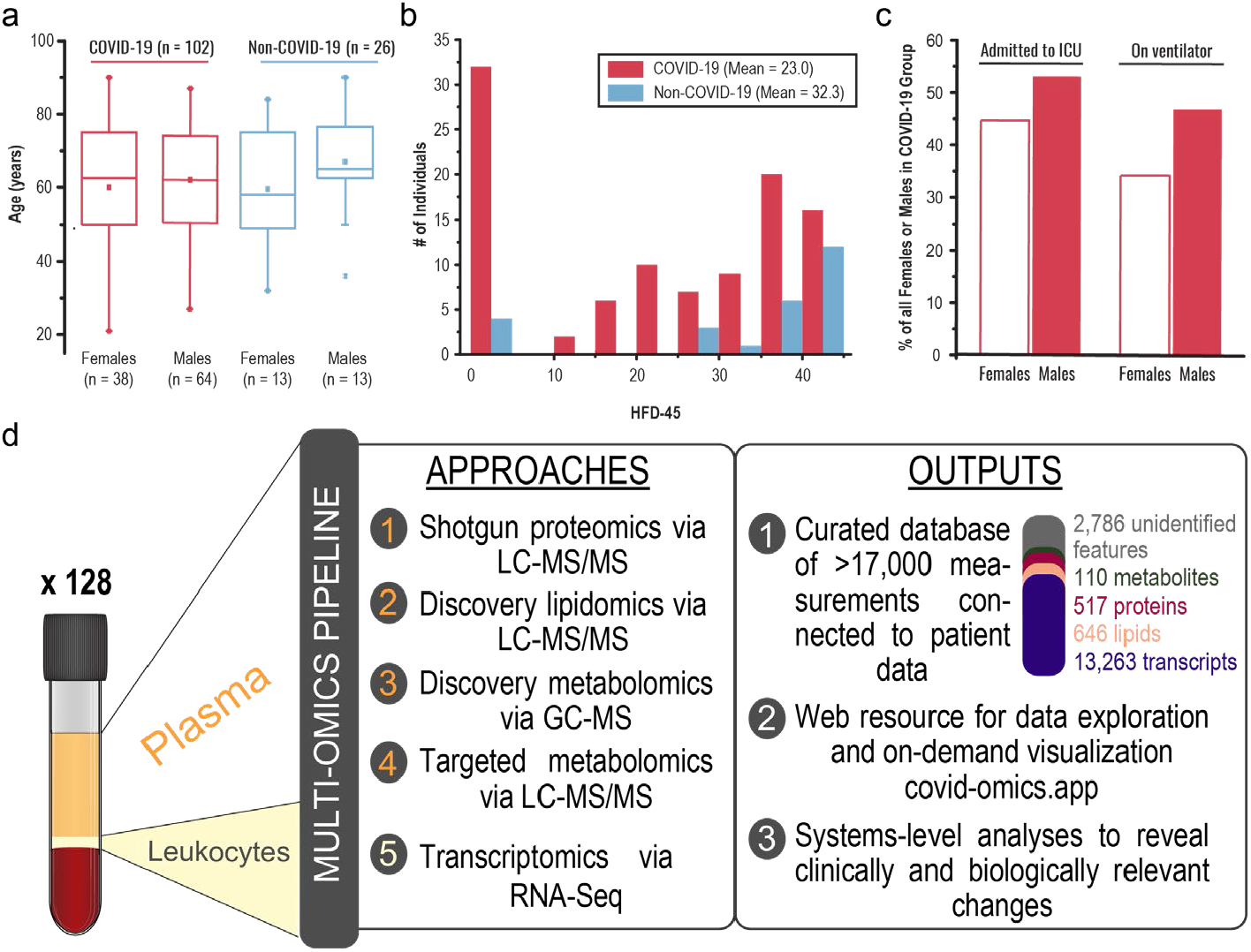
Overview of sample cohort and experimental design. **a** Age and sex distributions of COVID-19 (n = 102) and non-COVID-19 (n = 26) groups. **b** Distributions of hospital-free days over a continuous 45-day period aggregated with survival (HFD-45, see outcomes selection in the methods section) among COVID-19 and non-COVID-19 groups. **c** The proportion (%) of female and male patients who were admitted to the intensive care unit (ICU) and required support of a mechanical ventilator. **d** Overview of the study design, experimental approaches, and primary outcomes. Notice that the leukocytes were separated by filtering (see methods for details).

Severe COVID-19 infections require hospitalization and can lead to death due to respiratory deterioration. Thus, patients with the most severe cases requiring ventilatory support in the ICU entail longer admissions, while those with the most extreme cases die during hospitalization. To integrate length of hospitalization stay with mortality in one single outcome measure reflecting disease burden, we scored hospital-free days at day 45 (HFD-45). This composite outcome variable assigns a *zero* value (0-free days) to patients who remain admitted longer than 45 days or die while admitted and higher values of HFD-45 to patients with shorter hospitalizations and milder disease severity. Similar composite outcome variables are frequently used in the ICU setting <u>(Jaitovich et al., 2019; Young et al., 2015)</u>. Other clinically relevant information was obtained, including: Acute Physiologic Assessment and Chronic Health Evaluation (APACHE II) score, Sequential Organ Failure Assessment (SOFA) score, Charlson Comorbidity Index score, the number of days spent on mechanical ventilators, need for admission into the intensive care unit (ICU), and laboratory measurements of C-reactive protein (CRP), D-dimer, ferritin, lactate, procalcitonin, fibrinogen, and others (**Table 1**). Members of the non-COVID-19 group had on average considerably more HFD-45 than those of the COVID-19 group (32.3 vs. 22.0, respectively, p-value = 0.004; **Figure 1b**). APACHE II and SOFA severity scores, used to stratify severity of critical illness, were assigned to ICU-requiring patients, and both metrics exhibited similar distributions between the groups (**Table 1**). Consistent with previous reports, we observed male sex predominance in the ICU-requiring COVID-19 group (53.1 vs. 44.7%) and the group requiring mechanical ventilatory support (46.9 vs. 34.2%) (**Figure 1c**).

This comprehensive clinical characterization of the cohort provided substantial data to be integrated with multi-omic measurements. Following collection, blood samples were centrifuged to separate their components for various omic analyses (**Figure 1d**). We performed four individual mass spectrometry (MS)-based assays on the plasma components using high-resolution, high mass accuracy MS coupled with either gas or liquid chromatography: shotgun proteomics, discovery lipidomics, discovery metabolomics, and targeted metabolomics. RNA-Seq was used to characterize the transcriptomes of leukocytes extracted from the patient samples.

Collectively, these data enabled a comprehensive systems analysis of COVID-19 blood samples – containing abundance measurements of over 17,000 diverse biomolecules (517 proteins, 13,263 transcripts, 646 lipids, 110 metabolites, and 2,786 unidentified small molecule features). We compiled all biomolecule abundance measurements and de-identified patient information with clinical data into a highly curated relational (SQLite) database, which is publicly available for exploration and further analysis (see **Data Availability, Supplementary Figure 1**). The database includes additional metadata for each molecular measurement, such as gene ontology (GO) terms, alternative identifiers, and analytical metrics used to filter data, among others (see **Methods**). To facilitate exploration of this dataset by the broader scientific community, we also created a companion web tool (https://covid-omics.app) that accesses data and various analyses and performs on-demand data visualization (**Figure 1d**). Detailed description of this tool is presented in a later section.

### Systems analysis reveals strong biomolecule associations with COVID-19 status and severity

We hypothesized that measurement of a large number of biomolecules across different molecular classes (i.e., nucleic acids, proteins, lipids, and metabolites) would provide insight into the COVID-19 molecular landscape and facilitate understanding of factors associated with higher severity. To test this hypothesis in an unsupervised manner, we initially performed a principal component analysis (PCA) (**Figure 2a**). Here we note a strong grouping of patient samples based on severity (HFD-45 values) with samples of high and low severity diverging along the diagonal axis of principal components 1 and 2. This, together with a subtler grouping based on status (COVID-19 vs. non-COVID-19), merited further supervised exploration.

**Figure 2.**
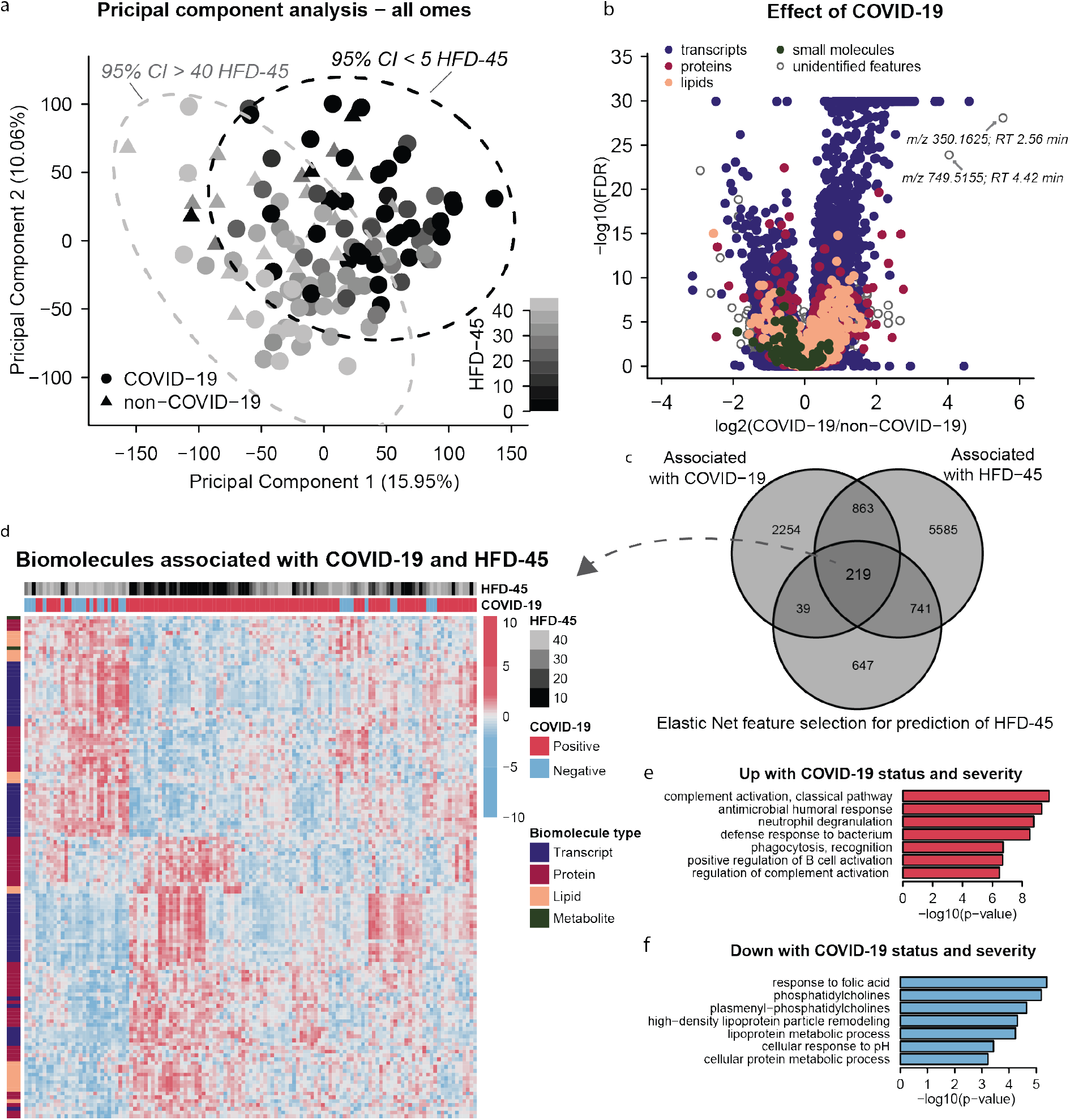
Multi-omics analysis reveals strong molecular signatures associated with COVID-19 status and severity. **a** Principal component analysis using quantitative values from all omics data (leukocyte transcripts, and plasma proteins, lipids, and small molecules) shows principal components 1 and 2 capture 16% and 10% of the variance between patient samples. Plotting samples by these two components show a linear tread with hospital free days at 45 days (HFD-45). **b** Associations of biomolecules with COVID-19 status was determined using differential expression analysis (EBseq) for transcripts, and linear regression log-likelihood tests for plasma biomolecules, the adjusted p-values (1 - posterior probability or Benjamini Hochberg-adjusted p _values_, respectively) are plotted relative to the log2 fold change of mean values between COVID and non-COVID samples. In total, 2,537 leukocyte transcripts, 146 plasma proteins, 168 plasma lipids, and 13 plasma metabolites had adjusted p-values < 0.05. **c** Associations between biomolecules and HFD-45 was estimated using a univariate linear regression (HFD-45 ∼ biomolecule abundance + age + sex) resulting in 7,408 biomolecules. A multivariate linear regression with elastic net penalty was applied to each omics dataset separately to further refine features of interest and resulted in 946 features. In total 219 features were determined as most important for distinguishing COVID status and severity. **d** The 219 features abundances were visualized via a heat map and clustered with hierarchical clustering. Features that were elevated (**e**) or reduced (**f**) with COVID status and severity were used for GO-term and molecular class enrichment analysis.

To gain biological insight into the host’s response to SARS-CoV-2 and pathways influencing its severity, we integrated our biomolecule measurements with clinical outcome variables. For this supervised analysis, we used univariate and multivariate regression to identify features which associate with (1) COVID-19 status and (2) HFD-45. Significant changes in plasma proteins, lipids, and metabolites associated with COVID-19 were determined by ANOVA and log-likelihood ratio tests, which incorporated potentially confounding variables, such as sex, age, and ICU status. Significant changes in leukocyte transcripts associated with COVID-19 diagnosis were analyzed separately by EBSeq. **Figure 2b** presents the statistical significance relative to the log_2_ of the fold changes of means for COVID-19 vs. non-COVID-19 groups for each biomolecule. In total, 2,537 leukocyte transcripts, 146 plasma proteins, 168 plasma lipids, and 13 plasma metabolites were significantly associated with COVID-19 status. Next, we applied GO and molecular class enrichment analysis to these features (**Supplementary Table 1**). Among the biological processes that were enriched in biomolecules whose abundance was altered with COVID-19 (combined proteins and transcripts), many support recent findings <u>(Messner et al., 2020) (Shen et al., 2020)</u>. These GO terms include mitotic cell cycle, phagocytosis recognition, positive recognition of B cell activation, complement activation (classical pathway), and innate immune response. Some of these enriched GO terms were driven by transcripts, for example mitotic cell cycle (38 transcripts vs. 0 proteins up with COVID-19), while other GO terms, like phagocytosis recognition or positive regulation of B cell activation, were driven by both transcripts and proteins (11 transcripts/18 proteins and 10 transcripts/18 proteins, respectively, **Supplementary Table 1**). Triacylglycerides (TGs) were also enriched and more abundant in COVID-19 samples, while plasmenyl–phosphatidylcholines (PCs) were less abundant in COVID-19 samples compared to non-COVID-19 samples. These results are in line with prior reports of significant changes in lipid composition in COVID-19 sera <u>(Shen et al., 2020)</u>.

In addition to the annotated features, 511 unidentified metabolites and lipids were also significantly associated with COVID-19 diagnosis. We performed additional spectral database searching and manually confirmed the identity of several of these features (**Supplementary Table 2**). Initially, we explored the features with the largest fold change between COVID-19 and non-COVID-19 (**Figure 2b**). The tandem mass spectrum from the unidentified feature with a mass-to-charge ratio (*m/z*) of 350.1625, and the largest fold-change in COVID-19 vs. non-COVID-19, contained a fragment ion diagnostic of chloroquine/hydroxychloroquine (*m/z* 247.10, C_14_H_16_ClN_2_) suggesting this top feature is a metabolite of the experimental therapeutic hydroxychloroquine (**Supplementary Figure 2**). The feature with the second largest increase in abundance in COVID-19-positive plasma (**Figure 2b**) having *m/z* value of 749.5155 was matched to the antibiotic Azithromycin (**Supplementary Figure 2**). By referencing patient medical records (**Table 1**), we determined that 85% of COVID-19 patients received hydroxychloroquine and 80% of COVID-19 patients received azithromycin treatment prior to sample collection (vs. 0% and 8% non-COVID-19 patients, respectively), thus explaining the strong association with COVID-19 status. This ability to detect drug metabolites highlights both the power of this discovery approach and the complexity of these samples, as these biomolecular data provide an archival-quality detailed snapshot of both the patients’ physiological response and exposures to therapies.

One ongoing challenge with COVID-19 disease is the broad and unpredictable range of patient outcomes, i.e., disease severity. To find biomolecules associated with severity, we regressed HFD-45 against abundance of each biomolecule. Using this univariate approach, we found 6,202 transcripts, 189 plasma proteins, 218 plasma lipids, and 35 plasma small molecules which were associated with disease severity. To further refine features of interest, for each molecule-type we performed multivariate linear regression on HFD-45 using the elastic net penalty <u>(Zou and Hastie, 2005)</u> (**Methods**). The elastic net simultaneously performs regression and feature selection; the elastic net penalty induces sparsity in the fit coefficients, which leads to selection of key features that best predict the response variable. In contrast to other approaches that also incorporate sparsity, such as LASSO, elastic net is known to have a “grouping effect” where it selects groups of correlated, predictive features enabling more informative results in downstream enrichment analyses. With the elastic net approach, 497 transcripts, 382 proteins, 140 lipids, and 60 metabolites were retained as predictive features for the outcome HFD-45. In total, we used the following criteria: (1) significance with COVID-19 status, (2) significance with HFD-45, and (3) elastic net feature selection, and generated a list of 219 features most significantly associated with COVID-19 status and severity (**Figure 2c** and **Supplementary Table 3**). Levels of these biomolecules in each sample are visualized in a heatmap in **Figure 2d**. Hierarchical clustering in the heatmap shows clear grouping by COVID-19 status and severity and reveals clusters of molecules (across omes) with similar trends across patient samples.

Applying GO term and molecular class enrichment analysis to these 219 most differentially abundant features (**Figure 2e-f**), we found that biomolecules in GO terms for complement activation (classical pathway), antimicrobial humoral response, and neutrophil degranulation were enriched with COVID-19 severity (**Supplementary Table 1**). The “complement activation” term encompassed many variable segments of immunoglobulins encoded by genes that undergo recombination to allow expression of light or heavy chains; twelve of them (8 IGHVs, 2 IGHVs, and 2 IGLVs) correlated with and predicted lower HFD-45 (**Supplementary Table 3**). In general, the complement system is involved in immune response to virus infection <u>(Stoermer and Morrison, 2011)</u> and has been postulated as a target for therapeutics <u>(Risitano et al., 2020)</u>. Our observations about immunoglobulins offer a glimpse into the early humoral response to SARS-CoV-2 and the relationship between preferential variable segment use and individual response trajectories. To better understand production of protective or possibly injurious antibodies, serial studies and characterization of the properties of individual antibodies is required <u>(Ju et al., 2020; Seydoux et al., 2020)</u>. Another top feature was pulmonary surfactant-associated protein B (SFTPB), notable because circulating SFTPB correlates with decreased lung function in smokers <u>(Leung et al., 2015)</u>, and may be a surrogate marker of lung deterioration in COVID-19 individuals.

Features that were reduced in COVID-19 samples and whose abundance was lowest in the most severe cases were enriched in the following categories: response to folic acid, PCs, plasmenyl-PCs, and high density lipoprotein particle remodeling. These features suggest a significant change in plasma lipid regulation. Similar to previous reports, we observed lower levels of APOA1, APOA2, and APOM in COVID-19 patients relative to non-COVID-19 patients <u>(Messner et al., 2020)</u>; <u>(Shen et al., 2020)</u>.

### Cross-ome correlation analysis

A major strength of multi-omic approaches is the ability to uncover functional connections between different classes of biomolecules. This effort is particularly important for small molecules, especially lipids, whose functions are often less well annotated. To reveal these connections, we integrated all omic measurements acquired on plasma samples by performing cross-ome correlation analysis. We calculated Kendall Tau coefficients in the pairwise fashion for all features. The heat map in **Figure 3a** reveals significant hierarchical clustering of Kendall Tau coefficients between proteins (rows) and small molecules (lipids and metabolites; columns) with statistical significance for their correlation with each other. Significance for COVID-19 status and HFD-45 is also colored as a binary metric where q-value <0.05. The approach identified many dense clusters with strong associations across biomolecule classes.

**Figure 3:**
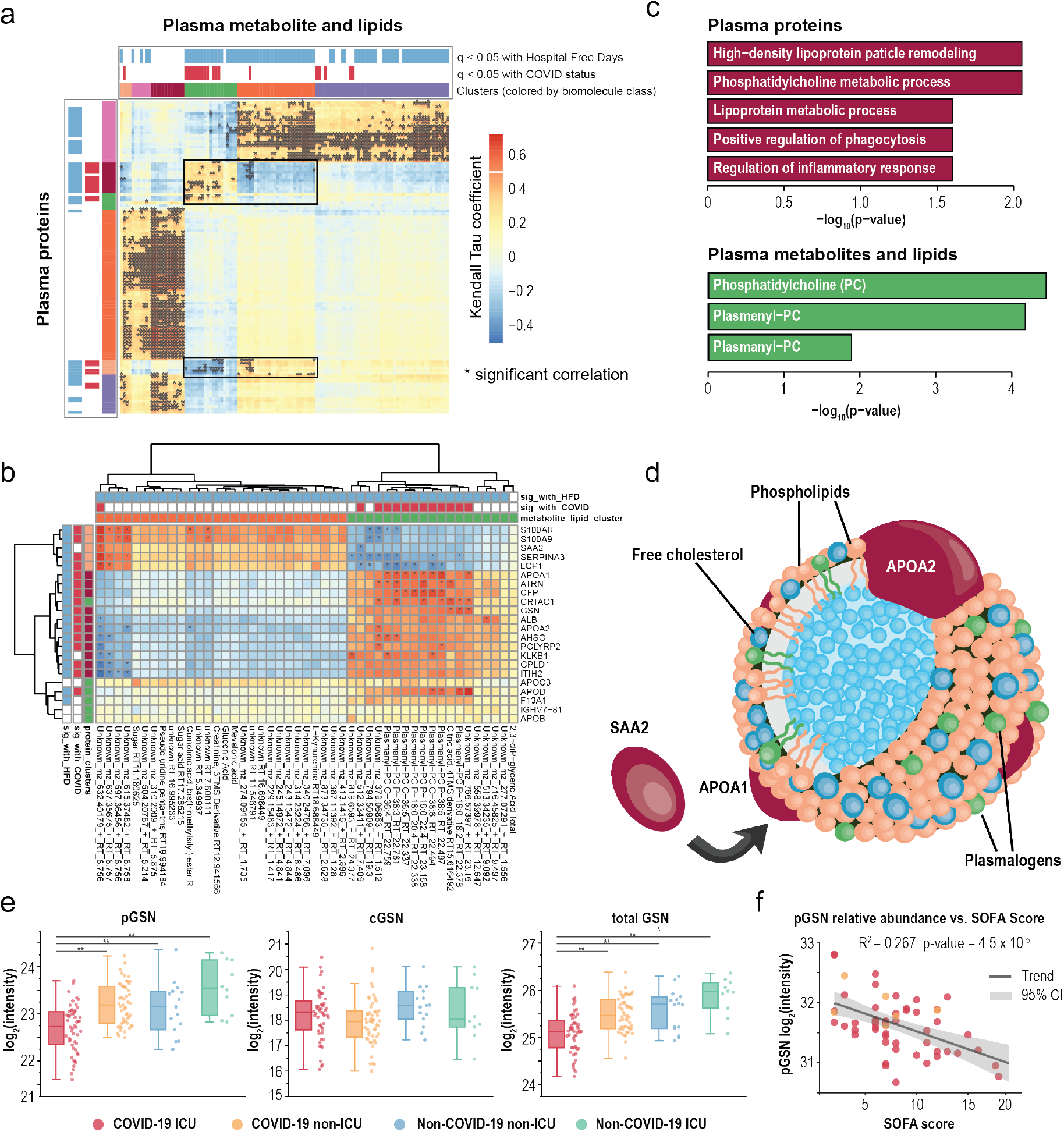
Leveraging the value of multi-omic data through cross-ome correlation analysis. **a** Hierarchical clustering of Kendall Tau coefficients calculated for correlations between abundances of proteins (rows) and small molecules (lipids and metabolites; columns) in the pairwise fashion. Significance of their association with HFD-45 and COVID-19 status is indicated above the biomolecule clusters. **b** Re-clustering of biomolecules found in the clusters highlighted in panel a with molecule annotations. **c** Enrichment analyses of protein GO terms (purple) and small molecule classes (green) present in the cluster in panel b. **d** A schematic of a high-density lipoprotein (HDL) particle containing APOA1 and APOA2 proteins surrounded by various lipids, specifically plasmalogens. SAA2, also detected in the cluster in panel b, can replaced APOA1 within the particle. **e** Relative abundance measurements of plasma gelsolin (pGSN), cellular gelsolin (cGSN), and total gelsolin obtained using parallel reaction monitoring (PRM) on representative peptide sequences. * and ** indicate p-values < 0.05 and 0.001, respectively. **f** Regression analysis of plasma gelsolin levels and SOFA scores (R^2^ = 0.267, p = 4.53 × 10-5).

One cluster with a diverse mix of biomolecules and multiple associations between each other and with COVID-19 status and severity is highlighted in **Figure 3b**. This cluster captured the lipids PC and plasmenyl-PCs and high density lipoproteins (**Figure 3c**). These categories were found to be significantly associated with COVID-19 status and severity (*vide supra*) and importantly this cross-ome analysis links these molecules as possible molecular interactors. We recently reported that plasma plasmenyl-PCs (containing 16:0) had strong genetic associations to the APOA2 locus in mice (Linke et al., in. press, http://LipidGenie.com), and now, with this cross-ome correlation analysis, we provide further evidence that these specific plasmalogen species are associated with APOA2 and APOA1 in humans. APOA1 and APOA2 are among the most abundant proteins in HDL particles (**Figure 3d**) <u>(Davidson et al., 2009)</u>. During proinflammatory response SAA2 can displace APOA1 within HDL particles and change HDL particles functions, and lead to increased clearance of HDL <u>(Feingold and Grunfeld, 2019; Khovidhunkit et al., 2004)</u>. Reduced levels of HDL are associated in increased risk of hospitalization for infections <u>(Trinder et al., 2020)</u> and poorer outcomes during infections <u>(Feingold and Grunfeld, 2019)</u>. The simultaneous reduction in apolipoproteins and plasmenyl PCs suggest that loss of these plasmenyl species also leads to poorer outcomes for COVID-19 patients. We suspect this may be due to the reduced capacity to handle oxidative damage, as plasmalogens are potent antioxidants <u>(Messias et al., 2018)</u>. These data highlight the potential of mitigating COVID-19 severity by using therapeutic strategies aimed at restoring HDL particles, potentially through supplementing plasmalogen lipids or balancing HDL and LDLs through statins <u>(Feingold and Grunfeld, 2019)</u>.

The notable cluster in **Figure 3c** also includes the protein gelsolin (GSN) and the metabolite citrate. Citrate is a calcium chelator that is used to prevent coagulation; intriguingly, GSN is a Ca^2+^-activated, actin-severing protein. Our cross-ome analysis offers the possibility that GSN and citrate are co-regulated in COVID-19 patients. Both features were among the 219 most significant features relating to COVID-19 infection and severity, and their reduced abundances were associated with worse outcomes. A possible confounding variable is that some patients received citrate as part of renal replacement therapy prior to sample collection; however, we find this confounder has no significant effect on the overall observation that citrate was lower in COVID-19 vs. non-COVID-19 (**Supplementary Figure 3**). The decrease in GSN in sera was recently reported by Messner et al., and confirming the reduced GSN levels in plasma samples (vs. sera) helps mitigate the confounding issue of variable loss of GSN into the fibrin clot <u>(Piktel et al., 2018)</u>.

Two proteoforms of GSN arise from the use of alternative transcriptional start sites: the cytoplasmic form, involved in remodeling of intracellular actin (cGSN) <u>(Feldt et al., 2019)</u>, and the plasma form, secreted by multiple cell types, including skeletal muscle (pGSN). Pools of circulating GSN, the aggregation of the two proteoforms, scavenge circulating filamentous actin and possess anti-inflammatory activities <u>(Piktel et al., 2018)</u>. As cGSN may enter plasma upon cellular injury, we used parallel reaction monitoring to independently quantify levels of pGSN and cGSN via targeting five distinguishing peptides (**Supplementary Table 4**) <u>(Peterson et al., 2012)</u>.

Abundance of pGSN correlated strongly with COVID-19 and ICU status, while cGSN levels were not reduced and did not correlate with COVID-19 severity (**Figure 3e**). These results indicate that cGSN from damaged cells is a relatively small contributor to the pool of circulating GSN and suggest that reduction of pGSN in COVID-19 may result from decreased biosynthesis and/or increased clearance from plasma.

Further, in the sub-group of ICU patients, decreased pGSN significantly correlated with SOFA scores (R^2^ = 0.27, p-value = 4.5 ⨯10^−5^, **Figure 3f**), demonstrating its potential utility as a prognostic biomarker of COVID-19 severity. These data also indicate that pGSN supplementation has excellent potential as a COVID-19 therapeutic. Indeed, pGSN is known to decrease in acute lung injury, and in animal models of lung injury repletion of pGSN has favorable effects<u>(Piktel et al., 2018)</u>. Recombinant pGSN has been given safely in phase I/II trials to patients with community-acquired pneumonia (NCT03466073, ClinicalTrials.gov). As of this writing, a trial of the recombinant pGSN in COVID-19 is being planned (NCT04358406).

### Biological processes dysregulated in COVID-19 patients

Based on recent literature reports on COVID-19 pathophysiology and our systems analyses, we next focused on specific biological processes evidently dysregulated in COVID-19 patients: neutrophil degranulation, vessel damage, platelet activation and degranulation, blood coagulation, and acute phase response.

### Neutrophil degranulation

Our list of 219 features strongly correlated to COVID-19 status and severity contained multiple transcripts and proteins involved in neutrophil degranulation (GO term 0043312; **Supplementary Table 1**). The volcano plot in **Figure 4a** illustrates the mean fold-change in abundance of GO-associated proteins (pink) and transcripts (purple), plotted against adjusted p-values of significance with COVID-19. We noticed an increased expression of multiple genes related to neutrophil function, including *PRTN3, LCN2, CD24, BPI, CTSG, DEFA1, DEFA4, MMP8*, and *MPO* (**Supplementary Table 3**). The latter encodes neutrophil myeloperoxidase, a protein instrumental in complexing extracellular DNA for the development of neutrophil extracellular traps (NETs) <u>(Jiao et al., 2020)</u>. NETs are extracellular aggregates of DNA, histones, toxic proteins, and oxidative enzymes released by neutrophils to control infections, and their overdrive can amplify tissue injury by inflammation and thrombosis <u>(Twaddell et al., 2019)</u>. NETs have been associated with the pathogenesis of ARDS <u>(Lefrançais et al., 2018; Middleton et al., 2020; Narasaraju et al., 2011)</u> and thrombosis <u>(Ali et al., 2019)</u>, both phenotypes observed in severe COVID-19 patients, and elevated neutrophil counts predict worse outcomes in COVID-19 patients <u>(Wang et al., 2020a)</u>. Interestingly, several NETs proteins that are also linked with thrombosis <u>(Healy et al., 2006; Wang et al., 2017)</u>, including calprotectin (S100A8/9), ferritin, CRP, and histone H3, were also increased in our cohort (**Table 1** and **Supplementary Table 3**). Thus our data suggest that combating dysregulated NETs may present an avenue towards mitigation of disease severity in COVID-19. For example, dipyridamole, an FDA-approved drug that can inhibit NET formation by activation of adenosine A_2A_ receptors <u>(Ali et al., 2019)</u>, has recently been shown to improve outcomes of COVID-19 patients with respiratory failure <u>(Liu et al., 2020)</u>. Similarly, the platelet P2Y12 receptor antagonist ticagrelor has been proposed to attenuate NET formation in COVID-19 <u>(Omarjee et al., 2020)</u> and may simultaneously inhibit platelet activation, a process also involved in COVID-19 pathophysiology <u>(Manne et al., 2020a)</u>. Drugs that antagonize the NET– IL1β loop, such as anakinra, are currently being tested (NCT04324021), and colchicine, a potent neutrophil recruiter and IL1β-secretion inhibitor, is also under evaluation (NCT04326790).

**Figure 4.**
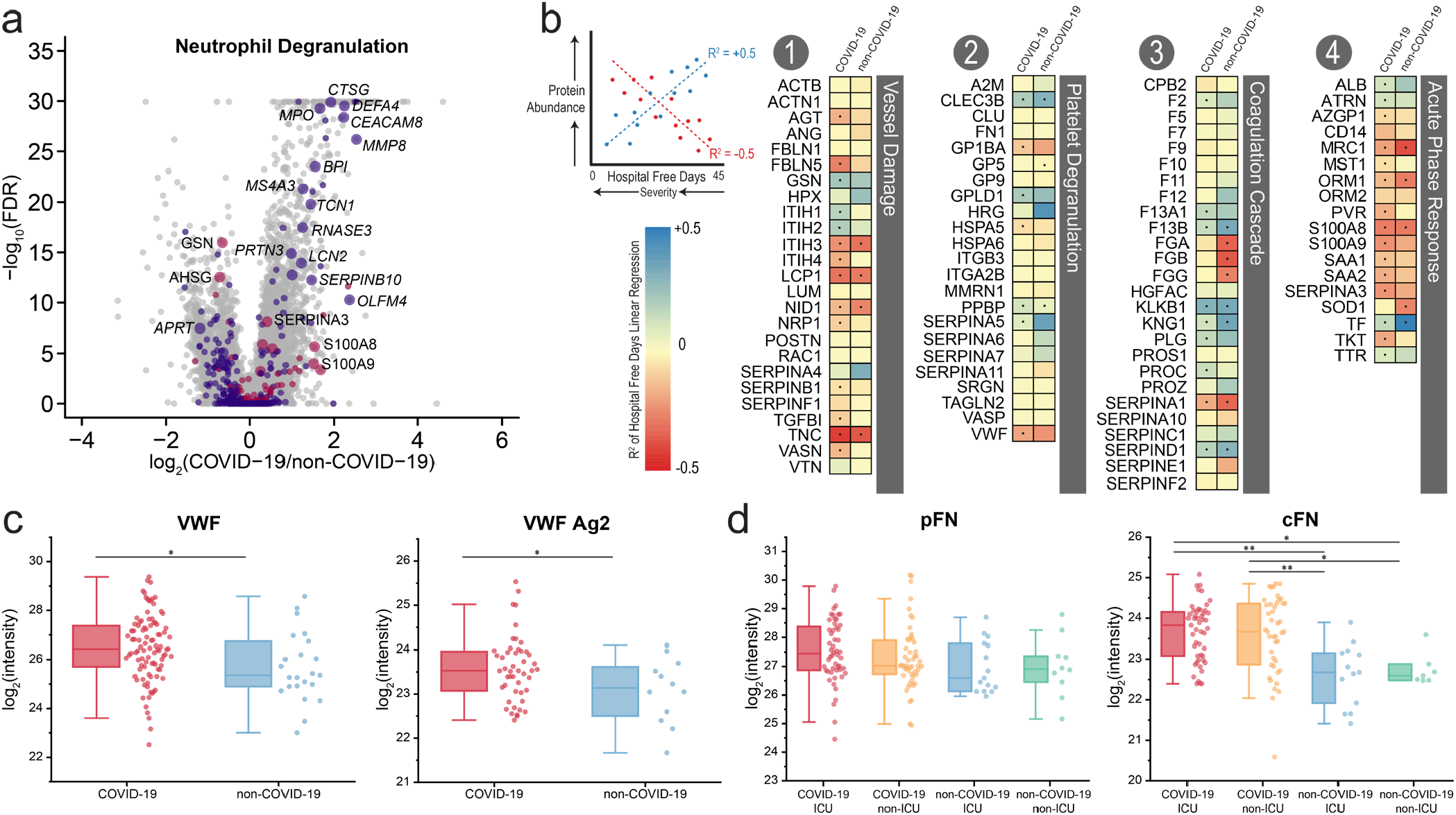
Biological processes dysregulated in COVID-19. **a** Volcano plots highlighting proteins (pink) and transcripts (purple) assigned with the GO term 0043312 “Neutrophil Degranulation.” Increased point size signified the inclusion of the biomolecule in the list of 219 features most significantly associated with COVID-19 status and severity (Figure 2e). **b** Linear regressions of protein abundance vs. HFD-45 for the indicated proteins as measured in COVID-19 (left) and non-COVID-19 patients (right). Resulting R2 values and their associated +/- slope indicate the goodness of fit and change in abundance of a given protein with severity (HFD-45). Proteins that are more decreased in severe cases appear blue, while proteins that are increased in severe cases appear red. Significance of the protein vs. HFD-45 correlation is denoted by a dot (p-value < 0.01). **c** Relative abundance measurements of peptides attributed to plasma fibronectin (pFN) and cellular fibronectin (cFN). **d** Relative abundance measurements of VWF multimer and VWF Antigen-2 (VWF Ag2), as estimated based on relative abundances of its unique peptides. Peptide- and protein-level data are log_2_-transformed and grouped into four categories, according to patient status: COVID-19 ICU (red), COVID-19 non-ICU (orange), non-COVID-19 ICU (blue), and non-COVID-19 non-ICU (green). * and ** indicate p-values < 0.05 and 0.001, respectively.

### Vessel damage

Numerous proteins involved in the body’s response to blood vessel damage, including AGT, FBLN5, NID1, and SERPINB1, increased in the COVID-19 samples relative to non-COVID-19 group and were particularly higher in abundance in more severe patients (**Figure 4b** and **Supplementary Figure 4**, *Category #1*). Neuropilin-1 (NRP1) is a regulator of the vascular endothelial growth factor (VEGF)-induced angiogenesis, and increase in its abundance with COVID-19 severity is of particular relevance given the recent report describing excessive pulmonary intussusceptive angiogenesis at autopsy in COVID-19 <u>(Ackermann et al., 2020)</u>. In contrast, other vessel damage-associated proteins, such as SERPINA4, significantly decreased in abundance in COVID-19 patients (p-value = 2.715 × 10^−4^), highlighting the regulatory intricacies of this biological process.

### Platelet degranulation

In accord with earlier reports, we observed severe dysregulation of proteins associated with platelet activation and degranulation (**Figure 4b**, *Category #2*) <u>(Manne et al., 2020b)</u>. For example, abundance of histidine-rich glycoprotein (HRG) - which binds heme, heparin, thrombospondin, and plasminogen <u>(Priebatsch et al., 2017)</u> - was significantly reduced in patients with COVID-19, with the mean level 1.8-fold lower than in non-COVID-19 patients (**Supplementary Figure 4**, p-value = 8.138 ⨯ 10^−12^). Similarly, abundances of GPLD1 and CLEC3B significantly decreased in COVID-19 and correlated with disease severity (**Figure 4b** *Category #2*, **Supplementary Figure 4)**. In contrast, the abundances of other platelet-associated proteins were increased in COVID-19 samples vs. non-COVID-19 samples (**Supplementary Figure 4**), for example, serglycin (SRGN) and the von Willebrand Factor (VWF); VWF has been recently implicated in COVID-19-associated endotheliopathy by the use of an FDA-approved antibody test <u>(Goshua et al., 2020)</u>.

VWF is synthesized by endothelial cells as a 2,813 amino acid-long protein that is processed intracellularly to form VWF antigen 2 (VWFAg2) and VWF multimer <u>(Haberichter, 2015)</u>. VWFAg2 is released constitutively into circulation, whereas VWF multimer is stored for later release upon stimulation of endothelial cells <u>(Nightingale and Cutler, 2013)</u>. To distinguish between these two products, we quantified 19 peptides unique to VWFAg2 and 107 peptides unique to VWF (**Figure 4c**). Abundances of both VWFAg2 and VWF multimer were increased and correlated with COVID-19 severity (p-values = 4.860 × 10^−2^ and 1.715 × 10^−2^, respectively). We propose that endothelial cells increase synthesis, package, and release both VWFAg2 and VWF multimers, particularly in severe COVID-19 cases.

Fibronectin (FN1) works alongside VWF and fibrinogen to mediate the interaction of platelets with endothelial surfaces of injured vessels <u>(Wang et al., 2014)</u>. Previous studies have reported increased FN1 levels in COVID-19 patients, relative to non-COVID-19 patients <u>(Stukalov et al., 2020)</u>, but did not distinguish cellular (cFN) and plasma (pFN) proteoforms. By monitoring unique peptides (IAWESPQGQVSR for cFN and ESVPISDTIIPAVPPPTDLR for pFN), we specifically determined the abundance of each and revealed that cFN was significantly increased (p-value = 2.8 ⨯ 10^−6^) in COVID-19 patients, relative to non-COVID-19 patients, whereas pFN was not (**Figure 4d**). Such a finding is of considerable interest given that forced over-expression of cFN in plasma of mice is associated with increased thrombo-inflammation <u>(Dhanesha et al., 2019)</u>.

### Coagulation cascade

Consistent with the widely reported hypercoagulative phenotype of COVID-19 patients <u>(Becker, 2020; Goshua et al., 2020; Lax et al., 2020)</u>, all our patients exhibited evidence of excessive clotting *in vivo* as demonstrated by the increased levels of circulating fibrin D-dimer (**Table 1**). To further examine the coagulative dysregulation in COVID-19 patients at molecular level, we deliberately conducted our proteomic analyses using plasma, which retains clotting factors otherwise depleted in serum. These plasma proteins, working alongside proteins of the vessel wall and platelets, undergo a cascade of regulated proteolytic reactions to generate thrombin, which converts fibrinogen to fibrin <u>(Kattula et al., 2017)</u>. We detected significant increases in abundance of fibrinogen alpha (FGA) and beta (FGB) in COVID-19 vs. non-COVID-19 patient plasma, which was consistent with the clinically measured Fibrinogen (**Supplementary Figure 4, Table 1**), but to our surprise gamma (FGG) chains showed less significant trends in COVID-19 patients. We also observed decreases in abundance of prothrombin (F2) and thrombin-activation factor XIII (F13A1, F13B) in COVID-19 samples compared to non-COVID, and these proteins were further decreased in the most severe patients (**Figure 4b** and **Supplementary Figure 4**, *Category #3*). Significant reductions and correlations with HFD-45 were found for plasminogen (PLG), the zymogen precursor of plasmin <u>(Urano et al., 2018)</u>; kallikrein (KLKB1) and kininogen (KNG1), which function in the intrinsic coagulation cascade leading to generation of thrombin <u>(Schmaier, 2016)</u>; heparin cofactor 2 (SERPIND1), which forms an inhibitory complex with thrombin <u>(Huntington, 2013)</u>; protein C (PROC), which is activated by thrombin to activated protein C (APC) <u>(Griffin et al., 2018)</u>; and protein C inhibitor (SERPINA5) (**Figure 4b, Supplementary Figure 4**). The decreases of abundance of PROC and SERPINA5 with severity assume greater significance when considered alongside the finding that soluble plasma thrombomodulin (THBD) rivals elevated VWF as a predictor of mortality in COVID-19 <u>(Goshua et al., 2020)</u>. THBD is normally tethered to the surface of endothelial cells, where it forms a complex with thrombin that efficiently activates PROC to APC <u>(Ikezoe, 2015)</u>. APC also serves to modulate inflammatory response, and administration of recombinant APC mutant has been proposed as a therapy to mitigate thromboinflammation which occurs with ARDS and in severe COVID-19 patients <u>(Griffin and Lyden, 2020)</u>.

Our data are pertinent to current discourse on prevention and treatment of the thrombotic diathesis in COVID-19. Several serpin-protease interactions, modified by heparin and heparin-like compounds, are being investigated as anti-coagulant therapeutics for COVID-19 patients <u>(Cattaneo et al., 2020)</u>. Our data suggest such compounds and alternative antithrombotic therapies (e.g., anti-platelets) may also be useful in the subset of most severely affected COVID-19 patients.

### Acute phase response

Next, we evaluated acute phase proteins – i.e., proteins whose plasma concentration changes by at least 25% during inflammatory disorders as a result of reprogramming by cytokines and are often secreted from the liver <u>(Gabay and Kushner, 1999)</u> – including S100A8, S100A9, SAA1 and SAA2. Our findings both support previous reports of significant increase in abundance of these four proteins in COVID-19 patient samples (p-values = 1.673 ⨯ 10^−3^, 1.966 ⨯ 10^−4^, 2.830 ⨯ 10^−2^, 9.71 ⨯ 10^−3^, respectively) and reveal their correlation with severity as estimated by HFD-45 (p-values = 1.408 ⨯ 10^−8^, 1.996 ⨯ 10^−7^, 1.808 ⨯ 10^−4^, 1.420 ⨯ 10^−5^, respectively) (**Supplementary Figure 4, Figure 4b** *Category #4*). We also found that attractin (ATRN), a protein involved in chemotactic activity <u>(Duke-Cohan et al., 1998)</u>, decreased in abundance COVID-19 samples vs. non-COVID-19 samples and was significantly associated with severity (p-value = 1.124 ⨯ 10^−3^). Four of these proteins were also found in the cross-ome correlation analysis (**Figure 3b**), where S100A8, S100A9, and SAA2 were negatively correlated with plasmalogens, while ATRN was positively correlated with these lipids.

Our data also revealed the increase in abundance of a liver-derived protein transketolase (TKT). One of the top 219 significant features (**Figure 2**), TKT was significantly elevated in COVID-19 patient plasma (p-value = 5.592 × 10^−4^, **Supplementary Table 3**). Other acute-phase proteins detected include GSN (*vide supra*) and SERPINA3, both of which decreased in abundance in COVID-19 patients and correlated with HFD-45 (p-value = 7.823 ⨯ 10^−4^ for SERPINA3). In general, abundances of most identified plasma proteins associated with the acute phase response were significantly altered with COVID-19 status and indicative of severity.

### An interactive web tool to facilitate COVID-19 multi-omic data analysis

A primary goal of this study was to create a multi-omic compendium of biomolecules and their abundances in COVID-19 that could aid in the elucidation of disease pathophysiology and therapeutic development. To this end we have developed a web-based tool with interactive visualizations that allows for simple and quick navigation of our resource and performs on-demand PCA, linear regression, differential expression, and hierarchical clustering analysis via a clustered heatmap (https://covid-omics.app, **Figure 5**). The webtool enables supervised and unsupervised, univariate and multivariate analysis of the omic datasets in the context of highly curated clinical data. While the web resource will enable global access without need for code development, the source code is openly available on GitHub (https://github.com/ijmiller2/COVID-19_Multi-Omics) and the underlying relational database and unprocessed data are freely available (**Methods**).

**Figure 5.**
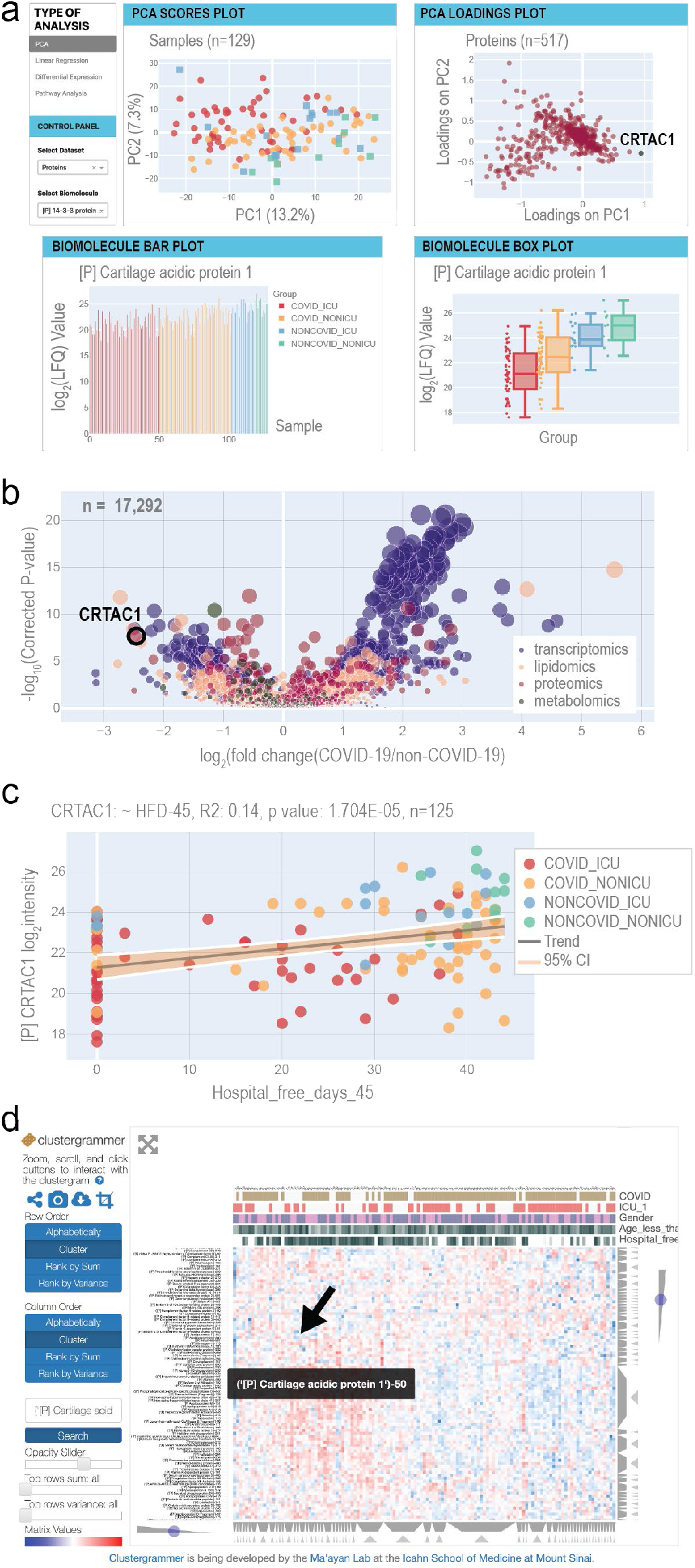
Overview of the COVID-19 Multi-omics Web Tool. **a** The home page provides principal component analysis (PCA) scores and loadings plots. Selected biomolecules are presented in a barplot and a boxplot. Each page provides buttons to navigate to the other web tools. **b** The differential expression page displays a multi-omics volcano plot with the y-axis representing -log10(p-values) where the p-values derive from the analysis in **Figure 2 c** The linear regression page allows users to select any combination of biomolecule and clinical measurement to analyze via univariate linear regression. R2 and p-values for the F-statistic are displayed on the plot. **d** The Clustergrammer page offers an interactive clustered heatmap.

To illustrate how this resource might be leveraged, we focus on a standout in the proteomics dataset, Cartilage acidic protein 1 (CRTAC1), which appears on the diagonal axis of the PCA loadings plot separated in both principal components 1 and 2 (**Figure 5a**). The biomolecule bar plots and boxplots, found just below the PCA plot on the resource’s homepage, show the per-sample abundance and distribution across sample groups of this protein, respectively (**Figure 5a**). Collectively, these data visualizations pinpoint CRTAC1 as an important feature in the innate structure of the proteomics data with a distinct abundance distribution for each patient subgroup (based on COVID-19 and ICU status), with the COVID-19 ICU group exhibiting the lowest average levels. CRTAC1 is a secreted matrix-associated protein involved in chondrocyte development and, interestingly, also belongs to the GO process “Olfactory Bulb Development” (GO:0021772), which is notable given previous reports of anosmia in COVID-19 patients <u>(Marinosci et al., 2020)</u>.

To learn how CRTAC1 relates to our clinical measurements, such as HFD-45, the user can redirect to the linear regression page - a flexible tool for generation of scatter plots and calculations of R^2^ values and (coefficient) p-values between any biomolecule and clinical covariate pair (**Figure 5b**). For the levels of CRTAC1 and HFD-45, the linear regression tool reports the R^2^ value as 0.14 and a p-value of 1.704e-05 (n=125). Checkboxes on the control panel of the page allow users to select various patient subgroups and independently calculate the metrics for them. For example, for the COVID-19 subset, the R^2^ is 0.079 and p-value is 4.490e-03 (n=100) (**Supplementary Figure 5a**), whereas for the non-COVID-19 subset of patient samples, the R^2^ is 0.099 and p-value is 1.254e-01 (n=25) (**Supplementary Figure 5b**).

The third analysis enabled by the web tool is differential expression. The page depicts a volcano plot, where effect size represents the log_2_ of the fold change of biomolecule abundance in COVID-19 vs. non-COVID-19 patient samples, and significance is -log_10_(p-value). Here the p-value is calculated using a likelihood ratio that compares linear models with and without possible confounders, including age, sex, and ICU status. For any given biomolecule, an accompanying table (not shown) lists the effect size, p-value, q-value (adjusted by false discovery rate [FDR]), type of statistical test, and associated formula used to calculate the p-values. This information provides users with a robust means of examining the implications of potential confounding variables. According to the differential expression analysis, abundance of CRTAC1 is the second most reduced in the comparison of COVID-19 vs. non-COVID-19 patients (**Figure 5c**). Even after accounting for age, sex, and ICU status, the adjusted p-value of the reduction in abundance of CRTAC1 in COVID-19 vs. non-COVID-19 samples is 2.236e-08.

The final data exploration tool is an interactive heatmap, powered by Clustergrammer <u>(Fernandez et al., 2017)</u> and provided to aid in investigation of how a biomolecule of interest may fit into larger patterns across samples (**Figure 5d**). Interestingly, CRTAC1 groups with a series of apolipoproteins (*vide supra*). Thus, as exemplified by our examination of CRTAC1, covid-omics.app permits facile and rapid exploration of any one of the >17,000 quantified biomolecules, providing a potential departure point towards novel biological insights.

### Two resource applications: comparative transcriptomics analysis and machine learning

To demonstrate how our resource could be leveraged by the scientific community, we present two complementary analyses: 1) we compare our transcriptomic data to the existing public transcriptomic datasets from patients with classical ARDS to elucidate gene expression signatures unique to COVID-19-induced ARDS, and 2) we utilize our multi-omic data to train machine learning models that accurately predict COVID-19 severity.

Severe COVID-19 is associated with development of ARDS, and although the evidence exists that COVID-19-induced ARDS is distinct from the classical ARDS <u>(Gattinoni et al., 2020b)</u>, more data are needed to clarify the differences between the two conditions. We performed differential expression analysis on transcriptomic data obtained on samples of COVID-19 patients fulfilling diagnostic criteria of ARDS in this study, *i*.*e*., patients in the ICU <u>(Ferguson et al., 2012)</u>, and an existing transcriptomic dataset on samples from patients with non-COVID-19 ARDS <u>(Englert et al., 2019a)</u>. After removing differentially expressed genes that were likely affected by batch effects (**Methods**), we found 436 and 470 genes more highly expressed in the COVID-19-ARDS and the non-COVID-ARDS cohorts, respectively (**Supplementary Figure 6**). We then examined these gene sets separately via the gene set enrichment analysis. Using an FDR threshold of 0.05, we found 30 gene sets to be enriched in the genes more highly expressed in the COVID-19-ARDS patients (**Figure 6a** and **Supplementary Table 5**). Among them was TNF-alpha signaling, involved in the cytokine storm of severe COVID-19 patients <u>(Catanzaro et al., 2020)</u>. TNF-alpha inhibitors have been proposed for treatment of COVID-19 <u>(Soy et al., 2020)</u>, and a TNF-alpha clinical trial has been registered <u>(ChiCTR2000030089)</u>. We also identified 79 gene sets enriched in genes more highly expressed in non-COVID-19-ARDS patients (**Figure 6b** and **Supplementary Table 5**). Several of them were related to viral response, *e*.*g*., decreased interferon signaling, suggesting that, in agreement with a recent report <u>(Blanco-Melo et al., 2020)</u>, COVID-19 may be characterized by a dysregulated leukocyte viral response. These results indicate that leukocytes from ARDS patients with and without COVID-19 possess overlapping, yet distinct transcriptomic profiles.

**Figure 6.**
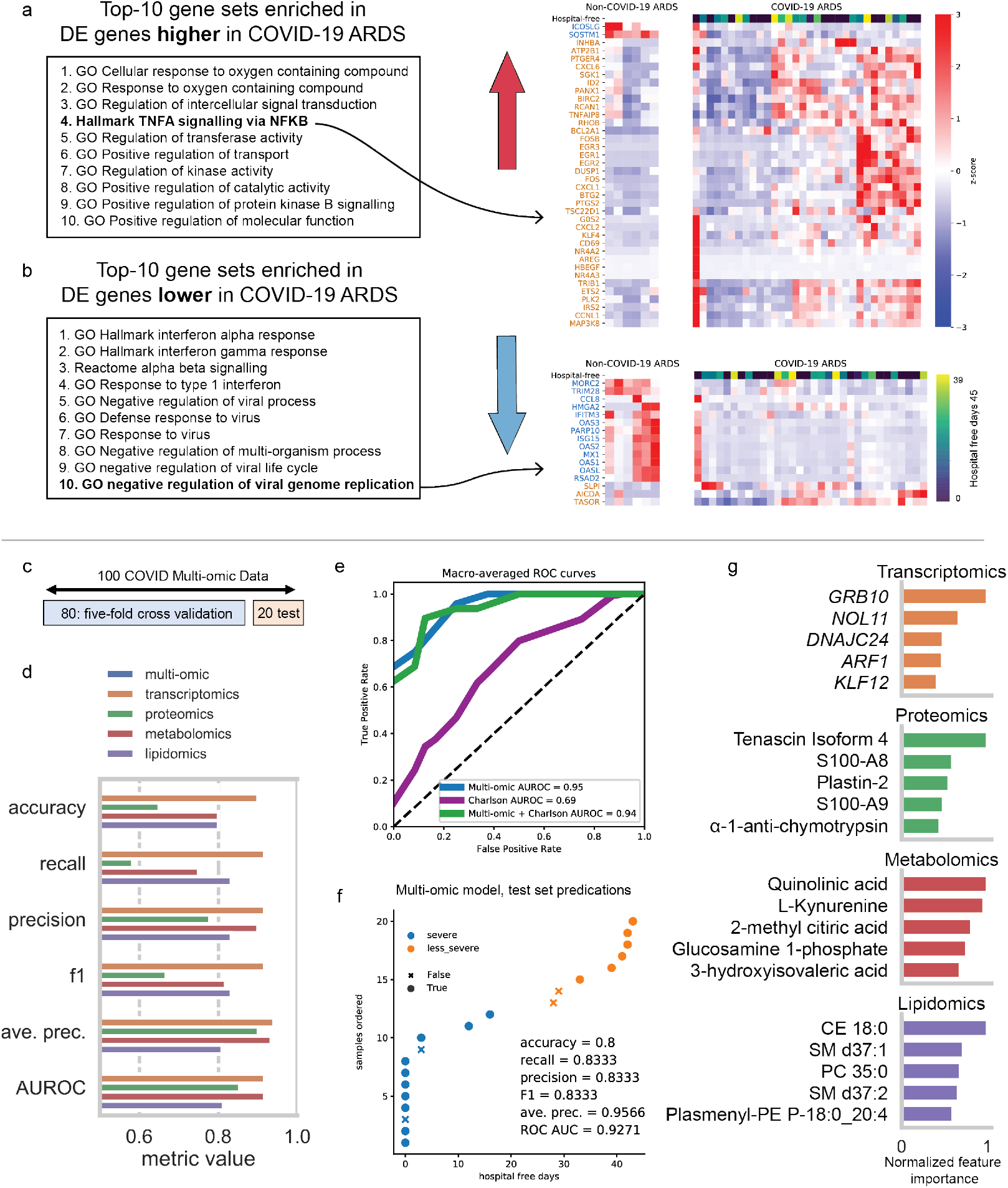
Results from analyses demonstrating use-cases of this multi-omic resource. **a** The top-ten enriched gene sets ranked by their adjusted p-value. For the gene set “TFNA signalling via NFKB”, we show a heatmap (right) of the z-score normalized expression data (in units of log transcripts per million) partitioned by whether the data came from the COVID-19-ARDS patients (right) or the non-COVID-19-ARDS (i.e. sepsis ARDS) patients from Englert et al. (left). The first row of each heatmap depicts the hospital-free days of each patient. We note that hospital-free days are not available for the Englert *et al*. dataset. The gene names labelling each row are colored according to whether the gene was deemed by EBSeq to be more highly expressed in COVID-19 ARDS (orange) or non-COVID-19 ARDS (blue). **b** Similar to (a); however, we instead analyze DE genes that are more lowly expressed in COVID-19 ICU patients. **c** Data splitting scheme for training and test sets from the 100 COVID patients with all four omic datasets. A random 20% was held out to be used for model evaluation, and the remaining 80% was used to determine the best hyperparameters with 5-fold cross validation. **d** Extra trees classifier performance metrics on the test set after hyperparameter optimization using each of the four omic datasets separately for training or all omic data combined. **e** Macro-averaged receiver-operator characteristic curves for the models trained with multi-omic data, Charlson score, or both multi-omic data and charlson score. **f** Test set predictions of the extra trees model trained on the combined multi-omic dataset showing correct predictions as a function of the disease severity defined by hospital free days. **G** Top 5 most important predictive features for each of the models trained on the four omic subsets. Feature importance for each set was normalized to the most important feature.

The ability to predict COVID-19 severity early in admission would help clinicians prioritize patients for treatments. Thus, we tested whether a machine learning model trained with the multi-omic data in our resource could accurately predict COVID-19 severity. The 100 COVID-19 patients with complete multi-omic profiles were assigned to “severe” and “less severe” groups based on HFD-45 using the median day 26 as the threshold. They were then randomly split into two groups: 80 were used for training and the remaining 20 were held out as a test set (**Figure 6c**). We optimized hyperparameters of ExtraTrees classification models using both the combined data and each of the four omic datasets separately via 5-fold cross-validation on the training set. This comparison shows that each omic dataset possessed different predictive utility (combined model AUROC = 0.93, average precision = 0.96; **Figure 6d**). Examination of the individual predictions from the combined model show that 2/4 incorrect predictions were for the patients with HFD-45 values near the median (**Figure 6e**), all of whom were also misclassified by the omic subset models (**Supplementary Figure 7**). Although preexisting comorbidities are associated with poor COVID-19 outcomes <u>(Guan et al., 2020)</u>, the model based on the combined multi-omic data was a substantially better predictor of disease severity than a model based on Charlson score, the clinical estimate of 10-year survival <u>(Richardson et al., 2020b)</u>. Adding Charlson score to the multi-omic data did not improve model performance (**Figure 6f**).

The use of the tree-based model also enabled determination of the most predictive features (**Figure 6g, Supplementary Table 6**). Some of them overlap with other analyses presented above, such as plasmalogen lipids and proteins S100A8 and S100A9. However, this model-based prioritization also highlighted potentially disease-relevant features not yet discussed. For example, for the metabolomic model, the most important features were tryptophan metabolites kynurenine and quinolinic acid, which are known mediators of immune function and inflammation in various contexts <u>(Heyes et al., 1992; Sofia et al., 2018)</u> and have been recently implicated in COVID-19 severity <u>(Migaud et al., 2020)</u>. Comparison of the top 25 predictive features from our proteomic and metabolomic models to those from another COVID-19 severity prediction model <u>(Shen et al., 2020)</u> revealed several overlapping features, including metabolite L-Kynurenine and proteins SAA2, SERPINA3, and LCP1. All in all, these results indicate that our multi-omic data can be utilized to accurately predict COVID-19 severity, and the feature importance from these models might further aid in understanding disease pathophysiology.

## DISCUSSION

In this cohort study, we used cutting-edge technologies to monitor thousands of diverse biomolecules from patients with and without COVID-19 in relation to the disease severity and outcomes. The overarching aim of this work was to capture the molecular signatures of COVID-19, correlate them with disease severity and clinical metadata, and generate both testable hypotheses and opportunities for therapeutic intervention. Altogether we make these data broadly available through a free web resource – covid-omics.app – with the hope that experts worldwide will continue to mine these data.

This resource has revealed several exciting findings that merit further exploration. First, our discovery that citrate, plasmenyl-PCs containing 16:0, and pGSN are covarying and reduced with COVID-19 severity is notable. Loss of these molecules would likely promote poorer outcome, and each has the potential to be supplemented in patients to mitigate the dysregulated response. pGSN, a circulating protein that regulates innate immunity by inhibiting proinflammatory mediators, inactivating bacterial endotoxin, and boosting endogenous antimicrobial peptides <u>(Piktel et al., 2018)</u>, is already being investigated as a therapy for treatment of community-acquired pneumonia and is an outstanding candidate for repurposing here. Citrate has been tested as a regional anti-coagulant in dialysis procedures and has also been noted to reduce negative effects of neutrophil degranulation <u>(Gritters et al., 2006)</u>. Monitoring and supplementing citrate levels could offer beneficial effects in severe COVID-19 patients. And lastly, treatment with statins or polyunsaturated lipid supplements could counteract the reductions in HDL in severe COVID-19 patients; statin use has already been associated with improved survival in COVID-19 patients <u>(Zhang et al., 2020b)</u>.

Our data both confirm the striking hypercoagulative signature of COVID-19 and expand on our current understanding of its pathophysiology. Anticoagulation strategies in non-COVID-19 critical illness have been extensively discussed, but no treatment has yet been identified. Evidence suggests that heparinization in COVID-19 patients may be beneficial <u>(Cattaneo et al., 2020)</u>, and mutant APC is another proposed therapeutic for the thrombo-inflammation of COVID-19 <u>(Griffin and Lyden, 2020)</u>. Our data offer several additional candidate therapeutics. For example, we observed significant reduction in abundance of prothrombin and its correlation with disease severity. Levels of another coagulation-related protein, cellular fibronectin (cFN), not pFN, were highly increased in COVID-19 patients. Finally, we detected dysregulation of VWF and circulating coagulation factors that could provide rationale for more tailored antithrombotic treatments, including the addition of several drugs synergistically working at different levels, e.g., coagulation and upstream endotheliopathy events.

Currently, the potential benefits of systemic corticosteroids in COVID-19 patients are under intense debate <u>(Huang et al., 2020; Wu et al., 2020)</u>. While patients with non-COVID-19 severe infections and ARDS represent an aggregation of different transcriptomic sub phenotypes <u>(Bos et al., 2017; Calfee et al., 2014; Davenport et al., 2016a)</u>, corticosteroids appear to be beneficial only to specific patient subgroups <u>(Antcliffe et al., 2019)</u>. Our RNA-Seq analysis will allow further characterization of immunomodulation targets that could be focused on the globally captured pathways, or on subpopulations associated with specific outcomes demonstrating enriched features. Moreover, the identification of a leukocyte and proteomic signature suggesting neutrophil extracellular trap formation (NETosis) could allow for the selection of already available drugs that target specific steps in neutrophil activation and function for COVID-19 repurposing.

The use of machine learning revealed additional features relevant to COVID-19 severity and underlined the utility of the multi-omics based model for predictions, as this model performed better than the well-established Charlson comorbidity index. Moreover, adding the Charlson score as a variable to our proposed model did not result in better predictive power. This finding could indicate that the clinical score is highly collinear with the multi-omic variables used by the model and that the clinical observation cannot fully capture the features leading to patients’ outcomes. Further, the combination of leukocyte transcripts with plasma biomolecules for predictive modeling provides a valuable resource for future investigations.

All large-scale omics studies have limitations but, ideally, still stimulate the generation of numerous testable hypotheses. This work is no exception; and, in a very real sense, it represents a starting point in the response to the urgent need to wholly define this devastating disease. Future work should include a larger and broader patient population. The samples derived here came from a single center, and although our population is racially diverse, it does not necessarily replicate factors related to geography or population socioeconomic status, among others. Another limitation was that the control group was generated by blindly enrolling patients presenting with COVID-19-compatible symptoms. While this approach provided an important reference, the non-COVID-19 patients admitted to the ICU, not all of these patients presented with the same criteria of ARDS. We recognize this approach is not ideal and expect that future studies with prospective enrollment of patients fully matching the present COVID-19 clinical features will provide a better reference to the current cohort.

The practicalities associated with study design and implementation during a pandemic presented another possible limitation. For example, while we enrolled the patients at the time of hospital admission, we could not control the period that elapsed between the disease development and the blood sampling. Nevertheless, previous research on the genomic landscape of patients with sepsis indicates the timing of blood sampling in relation to ICU admission was not predictive of the patients transcriptomic profiling <u>(Davenport et al., 2016a)</u>. Relatedly, we had no control of the various drugs administered to the patients based on recommendations including azithromycin, hydroxychloroquine, and others, which could have impacted the overall COVID-19 data generated. We note, however, that our detection of these therapeutics and their strong correlation with COVID-19 adds credibility to the quality of our analytical platform.

Disease burden was difficult to assess, and we relied upon the metric of HFD-45. Ultimately the data presented here conveys that this metric is a good outcome measure; however, our study was not powered to demonstrate the association of omic data with survival, which is the most impactful patient-centered outcome measure. Finally, our study relies on a single sample per patient, which, despite reflecting the patients’ status to some extent, is limited compared with trajectory analyses that could be contributed by serial blood sample determinations. Future studies will ideally have multiple temporal sampling timepoints per patient to allow for better controlled longitudinal severity correlations.

## Data Availability

Mass spectrometry raw files and the SQLite database file have been deposited to the MassIVE database (accession number MSV000085703). Raw fastq data and RSEM expression estimates are available at GEO accession XXXXXX (note, data have been submitted and we are waiting for an accession number assignment). RSEM expression estimates for Englert samples can be downloaded from the MassIVE database.

https://massive.ucsd.edu/ProteoSAFe/dataset.jsp?accession=MSV000085703

https://covid-omics.app

## ACKNOWLEDGEMENTS

We thank Christina Kendziorski, Robert Campbell, and Alicia Williams for helpful discussions. We also thank the Biotech Center core for the expedited RNAseq processing. This work was supported in part by the National Institutes of Health. Specifically, by the National Human Genome Research Institution through a training grant to the Genomic Sciences Training Program Grant 5T32HG002760 (T.M.P.), the National Heart, Lung, and Blood Institute (NHLBI) through awards K01-HL-130704 (A. Jaitovitch) and 5R01HL-049426 (H. A. Singer) and by the National Institute of General Medical Sciences through 1R01GM124133 (A. P. Adam) and The National Center for Quantitative Biology of Complex Systems (5P41GM108538, J.J. Coon). Further support was provided by the Collins Family Foundation Endowment (A. Jaitovitch), and the Morgridge Institute through a postdoctoral fellowship (M.N.B.).

## AUTHOR CONTRIBUTIONS

Conceptualization: K.A.O., E.S., I.M., J.B., M.J., A.A., B.S., H.S., R.S., J.J.C., A.J.; Methodology: K.A.O., E.S., J.B., B.P., A.J.; Validation: K.A.O., E.S., T.P.C., Q.Q., L.M., E.A.T., Y.H., B.P., Y.Z., L.S., V.L., D.M.; Software: I.M., D.R.B.; Formal analysis: K.A.O., E.S., I.M., J.B., M.N.B., T.P.C., J.M., E.A.T., Y.H., S.S., R.S.; Investigation: K.A.O., E.S., J.B., A.C., H.C., A.T., B.P., L.D., A.A., A.J.; Resources: B.S., H.S., R.S., J.J.C., A.J.; Data curation: K.A.O., I.M., J.B., D.R.B., S.S.; Writing - original draft: K.A.O., E.S., I.M., M.N.B., T.P.C., J.M., B.P., J.J.C., A.J. ; Writing - review/editing: K.A.O., E.S., I.M., J.B., M.N.B., T.P.C., J.M., Q.Q., E.A.T., D.R.B., L.S., A.A., B.S., H.S., S.S., D.M., R.S., J.J.C., A.J.; Visualization: K.A.O., E.S., I.M., J.B., M.N.B., T.P.C., J.M., L.M., D.R.B., Y.Z.; Supervision: K.A.O., E.S., I.M., J.B., R.S., J.J.C., A.J.

## METHODS

### Cohort characteristics

We conducted a single-center observational study of adult subjects admitted to either the medical floor or the medical intensive care unit (MICU) of Albany Medical Center. Ethical approval was obtained from the Albany Medical College Committee on Research Involving Human Subjects (IRB# 5670-20). Enrollment took place between April 6th, 2020 and May 1st, 2020 and follow-up continued until June 15, 2020. Patients were considered for enrollment if they were older than 18 years and were admitted to the hospital due to symptoms compatible with COVID-19 infection. Exclusion criteria were imminent death or inability to provide consent, which was obtained from the patient or a legally authorized representative. Patients who tested positive for COVID-19 were later assigned to that specific group and analyzed accordingly, and the COVID-19 negative group was composed of the remaining individuals. Prehospital comorbidities were determined by clinical history and with hospital documentation, and aggregated through the validated Charlson comorbidity index <u>(Charlson et al., 1987)</u>. Severity of critical illness at ICU admission was collected with the validated APACHE II and SOFA scores <u>(Ferreira et al., 2001)</u>. At enrollment, blood samples were collected in two plasma preparation tubes (PPT) tubes. One tube, used for Leukocyte RNA sequencing, was processed through LeukoLOCK® filters, and the remaining blood was centrifuged for plasma separation and aliquoted for further analysis. RNA was then eluted from LeukoLOCK filters following manufacturer recommendation, and samples were stored at -80 degrees celsius for later analyses.

### Selection of outcome measure

We intended to analyze the data with one encompassing outcome measure that fulfills the following criteria: 1) be able to combine severity of disease with mortality in one single metric; 2) be amenable to both ICU and medical floor populations; 3) use a timeframe that accounts for the fact that COVID-19 patients with respiratory failure require longer hospitalizations compared with non-COVID-19 individuals <u>(Wang et al., 2020b)</u>; (4) consider that COVID-19 causes a linear disease’s deterioration pattern that transition from mild respiratory compromise to respiratory failure, followed by respiratory distress requiring mechanical ventilatory support and eventually death. Thus, we selected the composite outcome variable “hospital-free days at day 45” (HFD-45), which assigns *zero* value to patients requiring admission longer than 45 days or who die during the admission, and progressively more free days depending on the hospitalization length.

### Plasma sample storage and aliquoting for mass spectrometry analysis

Samples were shipped on dry ice and kept in the -80°C freezer until analysis was performed. For MS acquisition, samples were randomly assigned into seven analytical batches. Samples for each batch were thawed on ice and mixed gently by tapping on a hard surface prior to aliquoting. In addition to patient plasma samples, an aliquot of pooled plasma sample (BioIVT, Human Plasma NaHep Lot# HMN378062) was extracted with each batch to serve as a batch quality control.

### Metabolomics

*Sample preparation:* 100 μL of plasma was aliquoted into 750 μL of Methyl Tert-Butyl Ether in a 1.5 mL Eppendorf tubes labelled with the sample name. This formed a biphasic separation, which was then vortexed for 1-2 s and shaken on an orbital mixer for 15 minutes. Then, 350 µL of a 1:1 chilled water:methanol mixture was added to each tube and then vortexed for 20 seconds and then centrifuged at 4 °C for 2 minutes at a speed of 14000 *x g* to separate the fractions completely. 250 μL of the bottom layer of this biphasic extraction was pipetted into a 1.5 mL Eppendorf tube containing 250 μL Acetonitrile to precipitate proteins. This mixture was centrifuged at 4 °C for 2 minutes at a speed of 14000 *x g* to pellet the protein completely. GC-MS: 100 µL of the extract was removed from the tube and collected into a low volume borosilicate glass autosampler vial with tapered insert. The volume of extract was dried in a vacuum concentrator until completely dry. The sample was resuspended in 50 µL methoxyamine hydrochloride solution (20mg/mL, pyridine solution), vortexed for 15 s and incubated at 37 °C for 90 minutes. Then, 100 µL of N-methyl-N-trimethylsilyltrifluoroacetamide (MSTFA) was added, vortexed for 15 s and heated at 60 °C for an additional 30 minutes. The sample was cooled at room temperature and then quickly centrifuged to ensure no sample remained near the vial’s cap. Samples were placed in the instrument autosampler at room temperature to await injection with no more than 24 hours between final centrifugation and analysis.

#### GC-MS

Samples were analyzed using a GC-MS instrument comprising a Trace 1310 GC coupled to a Q Exactive Orbitrap mass spectrometer (Thermo Scientific). A temperature gradient ranging from 50°C to 320°C was employed spanning a total runtime of 30 min. Analytes were injected onto a 30 m TraceGOLD TG-5SILMS column (Thermo Scientific) using a 1:25 split at a temperature of 275°C and ionized using electron ionization (EI). The mass spectrometer was operated in full scan mode using a resolution of 30,000. GC-MS raw files were processed using a software suite developed in-house that is available at https://github.com/coongroup. Following data acquisition, raw EI-GC/MS spectral data was deconvolved into chromatographic features and then grouped into features based on co-elution. Only features with at least 10 fragment ions and present in 33% of samples were kept. Feature groups from samples and background were compared, and only feature groups greater than 3-fold higher than background were retained. Compound identifications for the metabolites analyzed were assigned by comparing deconvolved high-resolution spectra against unit-resolution reference spectra present in the NIST 12 MS/EI library as well as to authentic standards run in-house. To calculate spectral similarity between experimental and reference spectra, a weighted dot product calculation was used. Metabolites lacking a confident identification were classified as “Unknown metabolites” and appended a unique identifier based on retention time. Peak heights of specified quant m/z were used to represent feature (metabolite) abundance. The data set was also processed through, where we applied a robust linear regression approach, rlm() function <u>(Marazzi et al., 1993)</u>, non-log_2_ transformed intensity values versus run order, to normalize for run order effects on signal.

#### AEX-LC-MS/MS

100 µL of protein precipitated solution (same as above for GC-MS analysis) was aliquoted into a low volume borosilicate glass autosampler vial with tapered insert. This volume was dried in a vacuum concentrator until completely dry. Then 100 µL of water was added to each tube to resuspend the contents and the sample was vortexed for 10 s to resuspend and then quickly centrifuged to ensure no sample remained near the vial’s cap. Samples were placed in the instrument’s autosampler at 4 °C to await injection with no more than 30 hours between final centrifugation and analysis. LC-MS/MS analyses were performed using a randomized sample order with a 10 µL injection volume. An Ultimate HPG-3400RS pump and WPS-3000RS autosampler cooled to 4 °C (Thermo Fisher) were mated to an IonPac AS-11 HC strong anion-exchange analytical column (2 mm ⨯ 250 mm, 4 µm particle diameter, Thermo Scientific) with AG-11 HC guard column (2 mm ⨯ 50 mm) heated to 40 °C. Mobile phase A (Waters) and mobile phase B (100 mM NaOH) were degassed and used in a gradient to elute analytes of interest. Column eluent passed through a 2 mm AERS 500e suppressor operated via a reagent-free controller (RFC-10, Dionex) set to 50 mA. The suppressor was operated in external water regeneration mode with water delivered at 0.5 mL/min by an Agilent 1100 binary pump. The multi-step gradient method was performed at 0.350 mL/min and performed as follows: 0.0 to 5.0 min (hold at 12.5% B), 5.0 to 10.0 min (12.5 - 20% B), 10.0 to 17.5 min (20 - 27.5% B), 17.5 to 22.5 min (27.5 - 42.5% B), 22.5 to 33.5 min(42.5 - 95% B), 33.5 to 34.0 min (hold at 95 % B, increase to 0.400 mL/min), 34.0 to 43.0 (hold at 95% B, 0.400 mL/min), 43.0 to 43.5 min (95 - 12.5% B, hold at 0.400 mL/min). The LC system was coupled to a TSQ Quantiva Triple Quadrupole mass spectrometer (Thermo Scientific) by a heated ESI source. The Ion Transfer Tube and Vaporizer Temp were kept at 350 °C. Sheath gas was set to 18 units, Auxiliary Gas to 4 units, and Sweep Gas to 1 unit. Both positive and negative spray voltage was static and set to 3,800 V. For targeted analysis the MS was operated in single reaction monitoring (SRM) mode acquiring scheduled, targeted scans to quantify selected metabolite transitions. All transitions and collision energies were previously optimized by infusion of each metabolite. The retention time window ranged from 3.5 min - 10 min and the “Use Calibrated RF Lens” option was selected. MS acquisition parameters were 0.7 FWHM resolution for Q1 and 1.2 FWHM for Q3, 0.8 s cycle time, 1.5 mTorr CID gas, and 30 s Chrom Filter. Raw files were processed using Xcalibur Quan Browser (v4.0.27.10, Thermo Scientific) with results exported and further processed using Microsoft Excel 2010. The prepared standard solution was used to locate appropriate peaks for peak area analysis.

### Shotgun proteomics

*Sample preparation*. 5 µl plasma from each sample were aliquoted into 1.5 ml Eppendorf tubes, containing 50 µl Lysis Buffer A (5.4 M guanidinium hydrochloride, 50 mM Tris, 10 mM TCEP, 40 mM chloroacetamide, pH ∼ 8). After brief, gentle mixing by inversion, tubes were placed into a sand bath and boiled at 100-110°C for 10 min. Samples were then allowed to cool down to room temperature, and 450 µl methanol (up to ∼90% vol/vol) were added. Proteins were precipitated by spinning at 13,000 g for 6 min, and supernatants were discarded. The tubes were briefly air dried, and protein pellets were resuspended in 100 µl Lysis Buffer B (8 M urea, 100 mM Tris, 10 mM TCEP, 40 mM chloroacetamide, pH ∼ 8). Samples were vortexed for ∼10 min to ensure the pellets dissolved completely. 7 µl LysC (1:50 enzyme: protein ratio; Wako Chemicals, Richmond, VA) was added to each sample; tubes were inverted to mix and incubated at 37°C for 1 hour. Then samples were diluted with 450 µl 100 mM Tris (pH=8.0), and 15 µl trypsin (1:50 enzyme: protein ratio; Promega, Madison, WI) was added. After 1 hr incubation at 37°C samples were acidified with 10% trifluoroacetic acid to final concentration of 1%. Digested peptides were extracted using Strata X SPE cartridges in a 96-well plate format. Samples were dried in a SpeedVac (Thermo Fisher, Waltham, MA) and resuspended in 0.2% formic acid. Peptide concentrations were determined using Quantitative Colorimetric Peptide Assay (Pierce, Rockford, IL), and 1 µg peptides were used in each analysis. *NanoLC-MS/MS*. A capillary column was fabricated in-house by filling self-pack PicoFrit 75–360 µm inner-outer diameter bare-fused silica shells with integrated 10 µm electrospray emitter tips (New Objective, Woburn, MA) with 1.7 µm, 130 Å pore size, Bridged Ethylene Hybrid (BEH) C18 particles (Waters, Milford, MA) to a final length of ∼40 cm using in-house built ultra-high-pressure column packing station (Shishkova et al. Analytical Chem 2018). The column was installed on a Dionex Ultimate 3000 nano HPLC system (Thermo Fisher, Sunnyvale, CA) and kept at 53°C inside an in-house made heater. The gradient was delivered using mobile phase A (0.2% formic acid in water) and mobile phase B (0.2% formic acid in 70% HPLC-MS grade acetonitrile) at a flow rate of 300 nl/min over a 90-minute gradient, including column wash and re-equilibration time. Electrosprayed peptide ions were analyzed on a quadrupole-ion trap-Orbitrap hybrid Eclipse® mass spectrometer (Thermo Scientific, San Jose, CA). Orbitrap survey scans were performed at a resolving power of 240,000 at 200 m/z with normalized AGC target of 250% ions and maximum injection time of 50 ms. The instrument was operated in the Top Speed mode with 1 s cycle time using an advanced precursor determination algorithm<u>(Hebert et al., 2018)</u> for monoisotopic precursor selection. Precursors were isolated in the quadrupole with an isolation window of 0.5 Th. Tandem MS scans were performed in the ion trap using turbo scan rate on precursors with 2-5 charge states using HCD fragmentation with normalized collision energy of 25 and dynamic exclusion of 10 s. Normalized ion trap MS/MS ion count target was set to 300% with the maximum injection time of 14 ms and the fixed m/z range of 150-1,350. *Data searching and processing*. Raw files were searched using MaxQuant quantitative software package <u>(Cox et al., 2014)</u> (version 1.6.10.43) against UniProt *Homo Sapiens* database (downloaded on 6.18.2019), containing protein isoforms and computationally predicted proteins. If not specified, default MaxQuant settings were used. LFQ quantification was performed using LFQ minimum ratio count of 1 and no MS/MS requirement for LFQ comparisons. iBAQ quantitation and “match between runs” were enabled with default settings. ITMS MS/MS tolerance was set to 0.35 Da. Lists of quantified protein groups were filtered to remove reverse identifications, potential contaminants, and proteins identified only by a modification site. LFQ abundance values were log_2_ transformed. Missing quantitative values were imputed by randomly drawing values from the left tail of the normal distribution of all measured protein abundance values <u>(Tyanova et al., 2016)</u>. Protein groups that contained more than 50% missing values were removed from final analyses. Relative standard deviations (RSDs) for each protein group quantified across all seven technical replicates of healthy plasma controls were calculated, and proteins with RSD greater than 30% were removed from final analyses.

### Parallel reaction monitoring (PRM)

Tryptic plasma peptides, generated for shotgun proteomics, were separated over 35 min gradients using the Dionex Ultimate 3000 nano HPLC system. Five peptide ions, whose sequences were representative of cellular gelsolin (cGSN), plasma GSN (pGSN) or shared between the two isoforms, were sequentially targeted for MS2 analysis using Orbitrap Eclipse for the length of five minute windows centered around their expected retention times (**Supplementary Table 4**). Precursor ions were isolated using a quadrupole with 1.3 Da isolation window and fragmented using HCD with an NCE of 25. Orbitrap MS2 scans of all fragments were performed over a 150-2,000 m/z scan range at a resolving power of 60,000 at m/z of 200 with an AGC target of 2×105 ions and maximum injection time of 350 ms. Identification and quantification of targeted peptides were performed using Skyline open access software package (version 20.1). 4-5 most intense and specific transitions were used to quantify peptide abundances, and area-under-the-curve measurements for each peptide were exported for further analysis.

### Lipidomics

#### Sample preparation

20 µL of plasma was extracted with 500 µL of ice-cold extraction solvent containing 3:1:1 n-Butanol:water:acetonitrile. Samples were vortexed for 10 s, incubated on ice for 5 min, vortexed for another 10 s, and then centrifuged at 14,000 × g for 2 min to precipitate protein. 200 µL of the supernatant was dried by vacuum concentration in amber glass autosampler vials and resuspended in 100 µL of 9:1 Methanol:Toluene. *Instrumental Analysis:* for LC-MS analysis, 10 μL of extract was injected by a Vanquish Split Sampler HT autosampler (Thermo Scientific) onto an Acquity CSH C18 column held at 50°C (100 mm × 2.1 mm × 1.7 μm particle size; Waters) using a Vanquish Binary Pump (200 μL/min flow rate; Thermo Scientific). Mobile phase A was 0.2% formic acid in water, and mobile phase B was 0.2% formic acid in IPA:ACN (90:10, v/v) with 10 mM ammonium formate. Mobile phase B was initially held at 5% for 1 min and then increased to 40% over 3 min. Mobile phase B was further increased to 70% over 11 min, then raised to 80.5% over 4 min, raised to 85% over next 2 min, raised to 89.5% over next 5 min, raised to 91% over next 3 min, and finally raised to 100% over 5 min and held at 100 % for 5 min. The column was re-equilibrated with mobile phase B at 5% for 9 min before the next injection. The LC system was coupled to a Q Exactive HF Orbitrap mass spectrometer through a heated electrospray ionization (HESI II) source (Thermo Scientific). Source conditions were as follows:

HESI II and capillary temperature at 275°C, sheath gas flow rate at 30 units, aux gas flow rate at 6 units, sweep gas flow rate at 0 units, spray voltage at |4.5 kV| for both positive and negative modes, and S-lens RF at 60.0 units. The MS was operated in a polarity switching mode acquiring positive and negative full MS and MS2 spectra (Top2) within the same injection. Acquisition parameters for full MS scans in both modes were 30,000 resolution, 1 × 10^6^ automatic gain control (AGC) target, 100 ms ion accumulation time (max IT), and 200 to 1600 m/z scan range. MS2 scans in both modes were then performed at 30,000 resolution, 1 × 10^5^ AGC target, 50 ms max IT, 1.0 m/z isolation window, stepped normalized collision energy (NCE) at 20, 30, 40, and a 30.0 s dynamic exclusion. *Data Processing*. The resulting LC–MS data were processed using Compound Discoverer 2.1 (Thermo Scientific) and LipiDex <u>(Hutchins et al., 2018)</u> (v. 1.1.0). All peaks between 1 min and 45 min retention time and 100 Da to 5000 Da MS1 precursor mass were grouped into distinct chromatographic profiles (i.e., compound groups) and aligned using a 10-ppm mass and 0.3 min retention time tolerance. Profiles not reaching a minimum peak intensity of 5×10^5, a maximum peak-width of 0.75, a signal-to-noise (S/N) ratio of 3, and a 3-fold intensity increase over blanks were excluded from further processing. MS/MS spectra were searched against an in-silico generated lipid spectral library containing 35,000 unique molecular compositions representing 48 distinct lipid classes (LipiDex library “LipiDex_HCD_Formic”, with a full range of acyl-chains included). Spectral matches with a dot product score greater than 500 and a reverse dot product score greater than 700 were retained for further analysis, with a minimum 75% spectral purity for designating fatty acid composition. Removed from the data set were adducts, class IDs greater than 3.5 median absolute retention time deviation (M.A.D. RT) of each other, and features found in less than 3 files. Data were additionally searched with Compound Discoverer 3.1 with the discovery metabolomics nodes for additional spectral matching to mzCloud and mzVault libraries but retaining the feature group and peak picking settings as detailed for the Compound Discoverer 2.1 analysis.

### RNA-Seq

Libraries were standardized to 2nM. Paired-end 2x50bp sequencing was performed, using standard SBS chemistry (v3) on an Illumina NovaSeq6000 sequencer. Images were analyzed using the standard Illumina Pipeline (version 1.8.2). For the Englert results, fastq files were downloaded from the Gene Expression Omnibus (GEO) (http://www.ncbi.nlm.nih.gov/geo/), accession GSE84439, samples/ runs SRR3923733-SRR3923739 (omitting SRR3923734) and SRR3923747-SRR3923753. All RNA transcripts were downloaded from the NCBI refseq ftp site (wget ftp://ftp.ncbi.nlm.nih.gov/refseq/H_sapiens/mRNA_Prot/*.rna.fna.gz). Only mRNA (accessions NM_xxxx and XM_xxxx) and rRNA (excluding 5.8S) was then extracted, and immunoglobulin transcripts were downloaded from ENSEMBL (IG_C, IG_D, IG_J and IG_V). We created a file mapping accession numbers to gene symbols, and then used rem-prepare-reference to build a bowtie-2 reference database.

Fastq files were trimmed and filtered using a custom algorithm tailored to improve quality scores and maximize retained reads in paired-end data. RNA-Seq expression estimation was performed by RSEM v 1.3.0 (parameters: seed-length=20, no-qualities, bowtie2-k=200, bowtie2-sensitivity-level=sensitive) <u>(Li and Dewey, 2011)</u>, with bowtie-2 (v 2.3.4.1) for the alignment step <u>(Langmead and Salzberg, 2012)</u>, using a custom hg38 reference. After the collation of expression estimates, hemoglobin transcripts were removed from further analysis, and TPM values were rescaled to once again total 1,000,000 in each sample. Differential Expression analysis was performed using the EBSeq package (v 1.26.0) <u>(Leng et al., 2013)</u> in R (v 3.6.2).

### Linear regression and calculation of log-likelihood ratios for determining significance with COVID-19

Log_2_ normalized abundance values were used as a response variable and the following models were fit against the data using the R statistical and graphing environment and lm() function (R version 3.3.3) <u>(Team and Others, 2017)</u>.

1. Normalized_abundance ∼ COVID status + Age + Sex + ICU status
2. Normalized_abundance ∼ Age + Sex + ICU status

Models (1) and (2) were compared using the anova() function in R, which returns the log-likelihood ratio and significance. The p.adjust() function was applied to resulting p-value tables using the ‘fdr’ method to return adjusted p-values.

### Linear regression and calculation of log-likelihood ratios for HFD-45 significance

HFD-45 were used as the response variable and the following models were fit against the data using the lm() function in R.

3. HFD-45 ∼ normalized_abundance + Age + Sex
4. HFD-45 ∼ Age + Sex

Models (3) and (4) were compared using the anova() function in R which returns the log-likelihood ratio and significance. The p.adjust() function was applied to resulting p-value tables using the ‘fdr’ method to return adjusted p-values.

### Elastic net regression analysis on HFD-45

we fit a least-squares, linear regression model with an elastic net penalty <u>(Zou and Hastie, 2005)</u> using each molecule’s value for all COVID-19 patients as covariates and HFD-45 as the response variable. For the transcriptomic data, we first transform the transcripts per million (TPM) output from RSEM using log(TPM+1). We also include age and gender as covariates. Specifically, elastic net solves the following optimization problem:

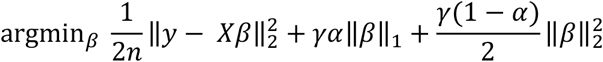

where n is the number of training samples, *X* is the matrix of covariates, *y* are the response variables, *X* are the model coefficients, *γ*is a parameter that sets the strength of regularization, and *γ*is a parameter that balances the strength of the L1 and L2 penalty-terms.

To determine the parameters for the elastic net model (i.e. and), we performed a grid search over a set of pairs of values for these two parameters where for each set of parameters, we perform five-fold cross-validation and evaluate each trained model according to the mean squared error on the held-out test set. We selected the best parameters according to this grid search and then used these parameters to fit the full dataset.

### GO biological processes and molecular class enrichment analysis

GO biological processes were obtained from Uniprot for transcripts and proteins. These terms were used, in addition to terms based on compound class (e.g., triglyceride), to perform a Fisher’s exact test for biomolecules within a subset relative to all measured biomolecules (phyper() function in R).

### Correlation analysis

Cor.test() function in R was used to perform cross-ome correlation analysis using method = Kendall. The Kendall correlation method is a rank-based approach and was chosen because it performs well with non-parametric data and, in contrast to Spearman, can perform better when values of the same rank occur <u>(Arndt et al., 1999)</u>.

### Comparison to transcriptomics data from Englert et al

Englert *et al*. RNA-Seq data were processed using the same computational pipeline used for our data. We examined all differentially expressed genes associated with an FDR value less than 0.01 and fold-change greater than two as determined by EBSeq <u>(Englert et al., 2019b; Leng et al., 2013)</u>. In total, this analysis produced 1,693 differentially expressed genes. Our RNA-Seq data originated from purified leukocytes, a method that can offer higher data quality<u>(Jiang et al., 2013)</u> and has recently been used to characterize the inflammatory transcriptomic phenotypes in sepsis <u>(Davenport et al., 2016b)</u>. This approach differed, however, from Englert *et al*. who generated data by sequencing RNA obtained from whole blood, and while they removed globin RNA, the difference in sample types could potentially contribute confounding batch effects. To mitigate this issue, we performed a second differential expression analysis between non-COVID-19 non-ARDS patients in our dataset and compared that with the non-ARDS sepsis control patients from Englert *et al*. We reasoned that genes that were similarly differentially expressed in both cohorts were likely to be genes attributed to batch effects. Thus, differentially expressed genes with FDR value < 0.05 produced by this overlapping analysis (a total of 787 of the 1,693) were removed.

### Gene set enrichment analysis for comparison with non-COVID-19 ARDS from Englert et al

We performed a separate gene set enrichment analysis on the 436 and 470 DE genes determined to be more highly expressed in COVID-19 ARDS and non-COVID-19 ARDS from Englert et al. (2019), respectively (**Supplementary Figure 6**). We considered all gene sets associated with Gene Ontology terms <u>(Ashburner et al., 2000)</u>, gene sets in the Hallmark collection <u>(Liberzon et al., 2015)</u>, and gene sets in the Canonical Pathway collection in the MSigDB repository <u>(Liberzon et al., 2011)</u>, which includes pathways from KEGG <u>(Kanehisa and Goto, 2000)</u> and Reactome <u>(Joshi-Tope et al., 2005)</u>. All gene sets were downloaded from MSigDB. To compute the enrichment of a given gene set with the DE genes, we used the GSEApy Python package (https://gseapy.readthedocs.io/en/latest/) to perform a Fisher’s Exact Test and adjusted all of the p-values using the Benjamini Hochberg Procedure <u>(Benjamini and Hochberg, 1995)</u>. We report all gene sets associated with an adjusted p-value less than 0.05 (**Supplementary Table 5**).

### Creation of covid-omics.app

The webtool was developed in Python (3.7.4) using the Plotly Dash package (1.12.0) and the source code is accessible via GitHub (https://github.com/ijmiller2/COVID-19_Multi-Omics/), under the src/dash directory. All package versions are described in the requirements.txt file. The R^2^ and p-value metrics displayed on the linear regression page are calculated using the statsmodels (0.11.1) library in Python. Note that, because the transcriptomics and lipidomics datasets have thousands of features, scatter plots in the web tool (including the volcano plot on the differential expression page and the loadings plot on the PCA page), only the top 1,000 features (ranked by variance across samples) are displayed to ensure reasonable speed and performance from the web server. However, all biomolecules that passed QC cutoffs are queryable using their respective drop down tools and available in the curated SQLite database (see **Data availability**).

### ExtraTrees classification of severe and non-severe cases

Molecular measurements of metabolites, lipids, proteins, and transcripts were read into Python from the SQLite database. Note unknown features detected in the metabolomic and lipidomic analyses were excluded from machine learning models. Patients were filtered to include only those with all four omic datasets, resulting in a total of 100 COVID-19 patients. 20% of the data was randomly held aside to be the true test set, and the other 80% of the data was used for 5-fold cross validation to determine the best model hyperparameters. Multiple classification models were compared with scikit-learn <u>(Pedregosa et al., 2011)</u>, and Extratrees Classification was found to perform best and used for further modeling. The threshold of HFD-45 to separate severe and non-severe cases was explored by training models with 5-fold cross validation at every threshold from 1-34 days. Multiple metrics were computed for each model, including accuracy, recall, precision, F1 score, average precision score, and receiver-operator characteristic area under the curve (AUROC or ROC AUC). The median HFD-45 of the 100 patients was chosen to split the patients into severe and less severe (26 days). ExtraTrees classifier hyperparameters were optimized separately for each omic data subset or the combined multi-omic data. Five folds were fitted for each of 5,940 candidates, totalling 29,700 model fits per dataset. A grid of the following parameters were tested: ‘n_estimators’: [100, 200, 300, 400, 500, 600, 700, 800, 900, 1000], ‘max_features’: [‘auto’, ‘sqrt’, ‘log2’], ‘max_depth’: [10, 31, 52, 73, 94, 115, 136, 157, 178, 200, None], ‘min_samples_split’: [2, 5, 10], ‘min_samples_leaf’: [1, 2, 4], ‘bootstrap’: [True, False]. All metrics were computed with the standard functions in scikit-learn, and the reported feature importances are the Gini type. All code is available on github as a jupyter notebook along with the python environment.yml for building the unique python environment with anaconda.

